# The Role of p16Ink4a as an Early Predictor of Physiological Decline during Natural Aging

**DOI:** 10.1101/2024.11.21.24317752

**Authors:** Lingzi Tang, Siarhei Hladyshau, Allison Ross, Kirsten A. Nyrop, Amy Entwistle, Hyman B. Muss, Natalia Mitin, Denis Tsygankov

**Author notes:** Corresponding authors: Denis Tsygankov.

## Abstract

Cellular senescence is a prominent accomplice of aging. The expression of gene p16ink4a has been established as a biomarker of cellular senescence in humans and animal models. However, it has not been extensively studied in clinical settings in the context of natural aging and the development of age-related diseases. Here, we report the results of a natural aging study that provided an assessment of cellular senescence and a battery of measures of clinical status, quality of life (QOL), and physical performance in 250 community-dwelling participants across age continuum. This report focused on analyzing predictive relationships between cellular senescence and different clinical assessments. Our results suggest that clinical labs and QOL assessments produce distinct groupings of participants, yet both have strong predictive associations with p16ink4a. Furthermore, the highest accuracy of p16ink4a prediction requires subsets of measurements representing diverse aspects of each assessment, pointing towards a system-level role of p16ink4a. Our analysis also led to an assessment-based composite indexes that strongly correlate with p16ink4a expression. Our study underscores p16ink4a’s association with both earlier signs of physiological decline (based on clinical labs) and the later onset of health issues limiting the quality of life.

## 1 INTRODUCTION

Life expectancy in the world’s population continues to increase, and globally, the population over 65 years is the fastest-growing group. In the U.S. alone, the population continues to age dramatically. By 2050, persons 65 and older in the U.S. will comprise about 90 million, more than double the same-aged population in 2010 [1]. However, these remarkable improvements in life expectancy have a downside: increased risk and frequency of diseases of aging [2] that result in disability, functional loss, poor quality of life, frailty, and high health care costs. Chronologic age alone is a poor indicator of biological age, as there is vast heterogeneity in health status within the same age groups. For older persons, high-quality tools are available that account for functional status, support, comorbidities, and nutrition and can be used to calculate life expectancy and disability accurately (https://eprognosis.ucsf.edu/index.php) [3]; no such tools exist for younger persons.

There is a significant need for accurate, biologically plausible, and easily accessible biomarkers that could predict aging trajectory, as well as risk and likelihood of illness in people who are considered “healthy”. The American Federation of Aging Research (AFAR) criteria for a valid biomarker of aging is one “that predicts a person’s physiological, cognitive, and physical function in an age-related way, must be testable and not harmful to test subjects, and should work in laboratory animals as well as humans” (www.afar.org; Biomarkers of Aging, 2016). p16Ink4a gene expression (further referred to as p16) emerges as a biomarker that can meet the AFAR criteria. Indeed, p16 is a cell cycle inhibitor and a widely recognized biomarker of cellular senescence that tends to increase with age [4]. Cellular senescence is a central biological process by which environment, genetics, and lifestyle affect human aging and lead to functional decline [5-13] [14, 15]. In murine models and almost all organs, p16 expression substantially increases in older mice compared to younger mice [16]. Senescent cells are characterized by permanent growth arrest, are metabolically active, and secrete numerous pro-inflammatory cytokines, contributing to inflammation, the development of diseases of aging [17], and further spread of senescence to healthy cells at both local and distant sites [12, 18]. Several genome-wide association studies implicate p16 as a critical determinant of human aging and age-related conditions [19-23].

In mouse models, the p16 promoter is used frequently to demonstrate the impact of cellular senescence on physiological decline and diseases. Injected and naturally occurring p16-positive cells have been shown to induce disease and shorten the healthy lifespan, while depleting p16-positive cells improves physical function and delays aging-associated disorders [5, 6, 12, 24, 25]. Recently, the p16 promotor has also been used to induce rejuvenation and improve health in mouse models by reprogramming of senescent cell states [26]. Given the prominent role of senescence in age-related decline, computational approaches based on machine learning models have been utilized in search of a reliable predictor of senescence, such as nuclear morphology [27, 28]. In human samples, p16 is also emerging as a common measure of cellular senescence across tissues [29].

With a strong indication of p16 being a biomarker of aging, here we sought to investigate if and how p16 correlates with early signs of physiological decline in a naturally aging population based on traditional clinical labs, quality-of-life surveys, and physical evaluation. To this end, we collected and analyzed data from 250 community-dwelling participants 25 through 85 years old. Our study showed that the expression of p16 in peripheral blood is significantly different between participants grouped based on both clinical labs and the RAND36 survey, although the clinical lab and QOL survey groupings do not fully overlap. The highest accuracy of p16 prediction requires diverse types of measurements within the different assessment categories, possibly indicating the systemic role of p16 in the physiological state. Finally, we derived linear-combination indexes for assessment categories that provide overall metrics strongly correlating with p16 expression (*r*∼0.35, which is over two-fold higher than any individual measurement).

## 2. METHODS

### 2.1 Study Participants

Participants were recruited from January to October 2022 (**Table 1 and Supplemental Figure 1**). Inclusion criteria: (1) age 25 to 85, (2) willing/able to attend all in-person visits and complete all study assessments and questionnaires, and (3) willing/able to provide written informed consent electronically. Exclusion criteria: (1) autoimmune disorders, (2) previous or currently undergoing chemotherapy, immunotherapy or radiation therapy for cancer, (3) history of transplants including solid organ or bone marrow, (4) presence of major active infection for which antibiotics and/or antivirals are prescribed within the last 14 days (chronic or acute, e.g., sepsis, HIV, pneumonia, active COVID infection), (5) dialysis, and/or (6) pregnancy. Recruitment was targeted by age cohort: N=30 aged 25-34, N=45 aged 35-44, N=50 aged 45-54, N=50 aged 55-64, N=45 aged 65-74, and N=30 aged 75-85. Recruitment was through word-of-mouth, Facebook, and the UNC Research for Me portal (researchforme.unc.edu). The study was approved by the Institutional Review Board of the University of North Carolina at Chapel Hill (IRB 21-2153).

**Table 1:** Participant characteristics. (statistics for demographics, comorbidities, clinical labs, and quality-of-life survey by gender are provided in **Supplemental Tables 1-2**)

| Characteristic | n (%) / mean ± std | Characteristic | n (%) |
| --- | --- | --- | --- |
| Age, mean ± std, years range | 54.3 ± 15.6, 25-85 | Diabetes or high blood sugar, n (%) | 14 (5.7) |
| Female gender, % | 181 (72.4) | Thyroid problems, n (%) | 30 (12.2) |
| Race: White, n (%) | 211 (84.4) | High blood pressure or hypertension, n (%) | 50 (20.3) |
| Race: Black or African American, n (%) | 10 (4.0) | Had a heart attack, n (%) | 7 (2.8) |
| Race: Asian, n (%) | 18 (7.2) | Stroke-like attack, n (%) | 4 (1.6) |
| Race: American Indian/Alaskan Native, n (%) | 1 (0.4) | Treated for heart failure, n (%) | 4 (1.6) |
| Race: More than one race, n (%) | 6 (2.4) | Emphysema, chronic bronchitis, or chronic obstructive lung disease, n (%) | 2 (0.8) |
| Race: Other, n (%) | 4 (1.6) | Asthma, n (%) | 42 (17.1) |
|  |  | Arthritis, n (%) | 74 (30.2) |
| Systolic BP, mean ± std, mmHg | 119.5 ± 17.6 | Stomach or peptic ulcer disease, n (%) | 14 (5.7) |
| Diastolic BP, mean ± std, mmHg | 78.0 ± 10.3 | Poor kidney function, used hemodialysis, or peritoneal dialysis, n (%) | 3 (1.2) |
| Heart rate, mean ± std, bpm | 66.8 ± 11.1 | Cirrhosis or severe liver damage, n (%) | 2 (0.8) |
| BMI, mean ± std, kg/m<sup>2</sup> | 26.0 ± 5.3 | Skin cancer, n (%) | 34 (13.8) |
| Average gait speed, mean ± std, m/sec | 3.1 ± 0.5 | Reproductive cancer, n (%) | 4 (1.6) |
| Up-And-Go time, mean ± std, seconds | 8.8 ± 1.9 | Non-reproductive cancer, n (%) | 3 (1.2) |

### 2.2. Questionnaires

Short Form SF-36 [30]. This 36-item questionnaire (further referred to as RAND36) inquired about general health, engaging in activities of daily living, problems in the past 4 weeks with work or other regular daily activities as a result of physical health or emotional problems, and general questions about fatigue, anxiety, or depression in the past 4 weeks. Response options are yes/no or multiple choice.

General health/lifestyle survey. Participants were asked whether living and/or deceased family members had a history of heart disease, cancer, diabetes, stroke, dementia/Alzheimer’s, COPD/pneumonia/flu, kidney disease, and accidents, and any family members who lived past 90 years of age. Participants were then asked if they had clinically diagnosed conditions such as diabetes or high blood sugar, thyroid problems, high blood pressure or hypertension, heart attack, stroke/cerebrovascular accident/blood clot or bleeding of the brain/transient ischemia attack, heart failure, emphysema/chronic bronchitis/chronic obstructive lung disease, asthma, arthritis, stomach ulcers/peptic ulcer disease, poor kidney function/hemodialysis/peritoneal disease, cirrhosis/severe liver damage, skin cancer, reproductive cancer, and/or non-reproductive cancer. Given that study enrollment occurred in early 2022, participants were also asked to list current medications, whether they had received a COVID-19 vaccine, and whether they had ever tested COVID positive.

### 2.3 Physical Evaluation

Timed Up & Go (UAG) [31]. For this test, participants stand up from a chair without assistance, walk 3 meters along a line on the floor at a normal pace, turn, walk back to the chair, and sit down. Patients who take more than 12 seconds to complete the test are considered at risk of falling.

Gait Speed [32]. Participants are in a standing position and are asked to walk at their normal speed: acceleration zone 1 meter, testing zone 4 meters, and deceleration zone 1 meter; then repeat.

Other health metrics: Blood pressure and heart rate were measured by a clinical coordinator using the same blood pressure cuff device (average of 3 measures). Body mass index (BMI) was calculated from height and weight measurements.

### 2.4 Clinical Labs

Blood samples were collected for Complete Blood Count (CBC), Comprehensive Metabolic profile (CMP), lipid panel, TSH, HbA1c testing. The complete registry is provided as **Supplemental Document 1**.

### 2.5 Gene Expression Assay

For p16 analysis, blood sample was collected into SapereX blood collection tube (provided by Sapere Bio) and T-cells were isolated within 72h.

mRNA expression in peripheral blood T-lymphocytes was determined using TaqMan real-time quantitative reverse transcription PCR. Expression analysis was performed by Sapere Bio (Research Triangle, NC), using technology described previously [33].

### 2.6 Gender considerations

The participant cohort consisted of 181 females and 69 males. To account for potential gender-related biases, we performed our analysis with and without normalization for gender. We chose not to use normalization based on the total average (or z-score) because gender differences may depend on age, and the subtraction of the global means (across all ages) may heighten gender differences for some of the measures in specific age groups. Instead, we performed the normalization based on linear regression. A running average or median could also be used, but would require introducing additional parameters (e.g., window size and sliding step for averaging), while simple linear regression does not. Specifically, all measurement values *V*_*M*_ for each male *i* were transformed to the normalized values 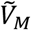 as

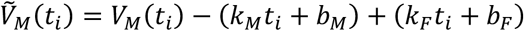

where *t*_*i*_ is the age of male *i*; *k*_*M*_ and *b*_*M*_ are the slope and intercept of the regression line relating the measure *V* and the age for all males (*V*_*M*_, *age*_*M*_); and *k*_*F*_ and *b*_*F*_ are the slope and intercept of the regression line relating this measure *V* and age for all females (*V*_*F*_, *age*_*F*_). We normalize males to females and not vice versa because females have about a 2.3-fold higher representation in our data. **Supplemental Figure 2** illustrates the normalization of the systolic blood pressure (SBP) as an example. Before the normalization, the regression slopes between SBP and age are slightly different for females and males. This difference is eliminated upon normalization. This example also illustrates that gender difference for SBP is higher in younger individuals, so aligning the all-age means would not eliminate the difference in SBP evenly for all ages, which could still bias our prediction models. Although some of the numerical results in our analysis end up varying between the results with and without normalization, our overall conclusions are not highly sensitive to this procedure. **Supplemental Tables 1-2** summarize the gender difference for all measures used in the following sections. The tables provide the *mean*, standard deviation (*std*), standard error (*se*), coefficient of correlation with age (*r*), p-value for testing the hypothesis of no correlation with age (*p*_*corr*_), and p-value from two-sample t-test for the hypothesis that the compared data comes from populations with unequal means (*p*_MF_). All numbers are provided for females and males with and without normalization. The color code highlights strong evidence for rejecting a null hypothesis (red: p-value < 0.05), weak evidence (yellow: p-value < 0.1), and insufficient evidence (green: p-value ≥ 0.1). The tables show that many measures have significant gender differences with respect to the correlation with age. For example, the thyroid-stimulating hormone (TSH) has a strong correlation with age in females (*r* = 0.23, *p*_*corr*_ = 0.002) but a weak one in males (*r* = 0.07, *p*_*corr*_ = 0.55), while the relative neutrophil count (Neut) has a weak correlation with age in females (*r* = 0.08, *p*_*corr*_ = 0.30) but a strong one in males (*r* = 0.38, *p*_*corr*_ = 0.001). The difference in the overall mean values is also significantly different for many measures before normalization. Expectedly, after our regression-based normalization, the difference in the overall mean becomes insignificant (*p*_*MF*_ > 0.1) for all measures.

### 2.7. Assessing the effect of age on pairwise correlations

A similar regression-based approach can be used to “age-correct” the data for each male *i* and female *j* and establish if there is a direct correlation between any two measures as opposed to an indirect correlation due to their common dependence on age:

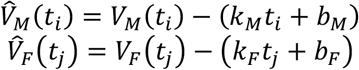

These age-corrected values for a given person can be viewed as measurement deviations from values typical for the person’s age. We considered three different scenarios:

Case 1: A pair of gender-normalized measures has a strong correlation (*p*_*corr*_ < 0.05) before age correction but no correlation (*p*_*corr*_ > 0.1) after age correction.

Case 2: A pair of gender-normalized measures has no correlation (*p*_*corr*_ > 0.1) before age correction but has a strong correlation (*p*_*corr*_ < 0.05) after age correction.

Case 3: A pair of gender-normalized measures has a strong correlation (*p*_*corr*_ < 0.05) both before and after age correction.

Because this work primarily focuses on the biomarker of cellular senescence, we present the results of these three case scenarios only for correlations between p16 and all 86 other measures from Supplemental Tables 1-2 (**Figure 1**).

**Figure 1.**
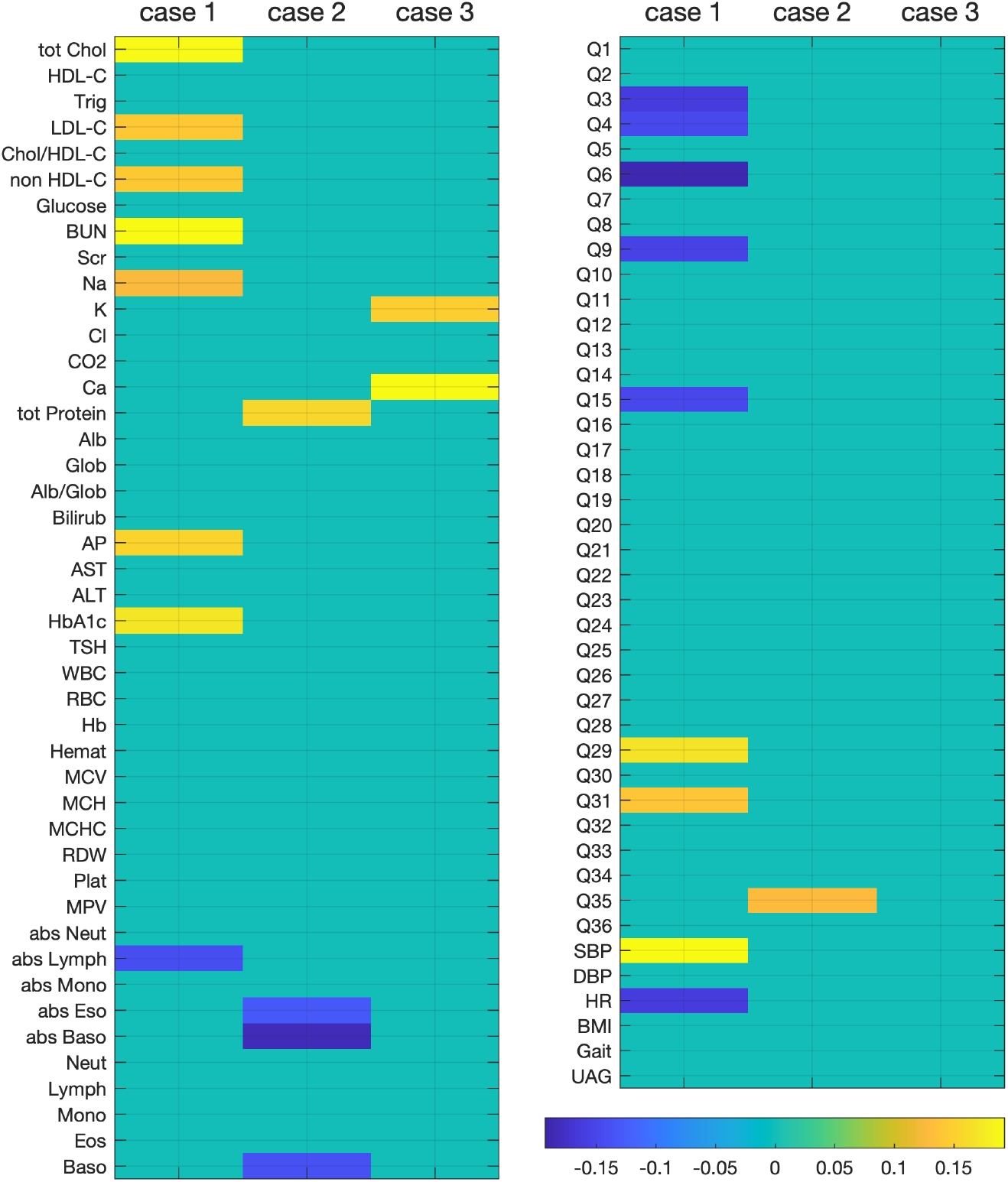
Comparison of the correlation coefficients, *r*, between p16 expression and all the other measurements. (left panel for clinical labs and right panel for RAND36 survey and physical evaluation). The correlation coefficients are shown for gender normalized data in three cases: **1)** when *p*_*MiFF*_ values less than 0.05 before age-correction and more than 0.10 after age-correction; **2)** when *p*_*MiFF*_ values are more than 0.10 before age-correction and less than 0.05 after age-correction; **3)** when *p*_*MiFF*_ values are less than 0.05 both before and after age-correction. In all cases, age correction is achieved by subtracting the regression lines in the measure vs age coordinates separately for male and female populations. Here, *p*_*MiFF*_ values validate the null hypothesis that there is no correlation between the measurements. *r* and *p*_*MiFF*_ values for all measurements before are after gender normalization are provided in **Supplemental Tables 1-2**.

In Case 1 (when *p*_*corr*_ values are less than 0.05 before age correction and more than 0.10 after age correction), p16 has a strong *positive* correlation with total cholesterol (tot Chol), LDL-C, non HDL-C, BUN, Na, alkaline phosphatase (AP), HbA1c, systolic blood pressure (SBP), and questions 29 and 31 on the RAND36 survey; and a strong *negative* correlation with absolute lymphocyte counts, heart rate (HR), and questions 3, 4, 6, 9, and 15 on RAND36 survey.

In Case 2 (when *p*_*corr*_ values are more than 0.1 before age correction and less than 0.05 after age correction), p16 has a strong *positive* correlation with total protein and question 35 on the RAND36 survey; and a strong *negative* correlation with abs Eso, abs Baso, and Baso.

In Case 3 (when *p*_*corr*_ values are less than 0.05 both before and after age correction), p16 has a strong *positive* correlation only with K and Ca.

These results show that very few individual clinical or QOL metrics still correlate with p16 when corrected for age (2/86 variables in Case 3 vs 17/86 in Case 1). Therefore, p16’s association with clinical labs, RAND36 survey, and physical evaluation is likely mediated by their common relationship with aging-related processes but not necessarily due to a direct dependence. Also, p16’s strong positive correlation with the total protein and Q35 on RAND36, as well as its strong negative correlation with absolute eosinophil and relative and absolute basophil counts become evident only after age correction. Thus, for these measures, p16 correlates not with the absolute values but with the extent of deviations from the mean values at that age.

### 2.8 Validation of hierarchical clustering method in application to the natural aging data

Pairwise correlations between the measures are informative but do not capture the higher-level interrelationships in the dataset. To investigate such data structures, we use both unsupervised clustering and hold-out training models. Before applying hierarchical clustering to grouping the data, we tested if our data had sufficient power to allow for meaningful multivariable associations. As the basis for determining if our data had sufficient power, we used the well-established 8-scale scoring of the RAND36 quality-of-life survey. This scoring system groups 36 questions into eight categories (health concepts): physical functioning, bodily pain, role limitations due to physical health problems, role limitations due to personal or emotional problems, emotional well-being, social functioning, energy/fatigue, and general health perceptions. Here, we test how accurately the hierarchical clustering algorithm can reproduce this grouping with our data from all 250 participants. **Supplemental Figure 3** shows a dendrogram for 36 survey questions. To provide a quantitative measure for the strength of the association of each question with each health concept, we applied the following method.

First, we selected one question from each concept as a landmark. For the landmarks, we used the questions that seeded the grouping (i.e., had the smallest dissimilarity with another question in the groups of the dendrogram shown in **Supplemental Figure 3**). Next, we randomly picked 90% of the participants, built a dendrogram for this subset, and measured Euclidian distances from each question to each landmark. Then, we repeated this procedure 5000 times (every time having a different random 90% sample) and averaged the result. Based on these averaged measurements, we found the distance to the closest landmark, *y*_*min*_, and to the next closest landmark, *y*_*next*_. This way, we not only associated each question with one of the eight landmarks but also determined a measure of the strength,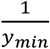, and exclusiveness, 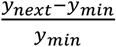, of this association. The second metric identifies cases when a question has an equally strong association with more than one health concept. **Figure 2** shows the resulting grouping and the corresponding metrics of the association strength and exclusiveness. For example, Q35 was misclassified as belonging to energy/fatigue instead of general health perception, but its exclusiveness metric is very 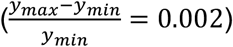, indicating that this question is on the borderline between the two concepts. Indeed, question 35, “I expect my health to get worse,” might reflect a possibility that the worsening prediction is significantly affected by the respondent’s low level of energy or high fatigue. The other two questions are Q30, “Have you been a happy person?” and Q3, “Does your health now limit you in vigorous activities, such as running, lifting heavy objects, participating in strenuous sports?”, may also fit well to “Energy/fatigue” instead of “Emotional well-being” or “Bodily Pain” instead of “Physical functioning.” In any case, our overall result is that only 3 out of 36 questions were misclassified with respect to the previously suggested grouping (91.7% accuracy).

**Figure 2.**
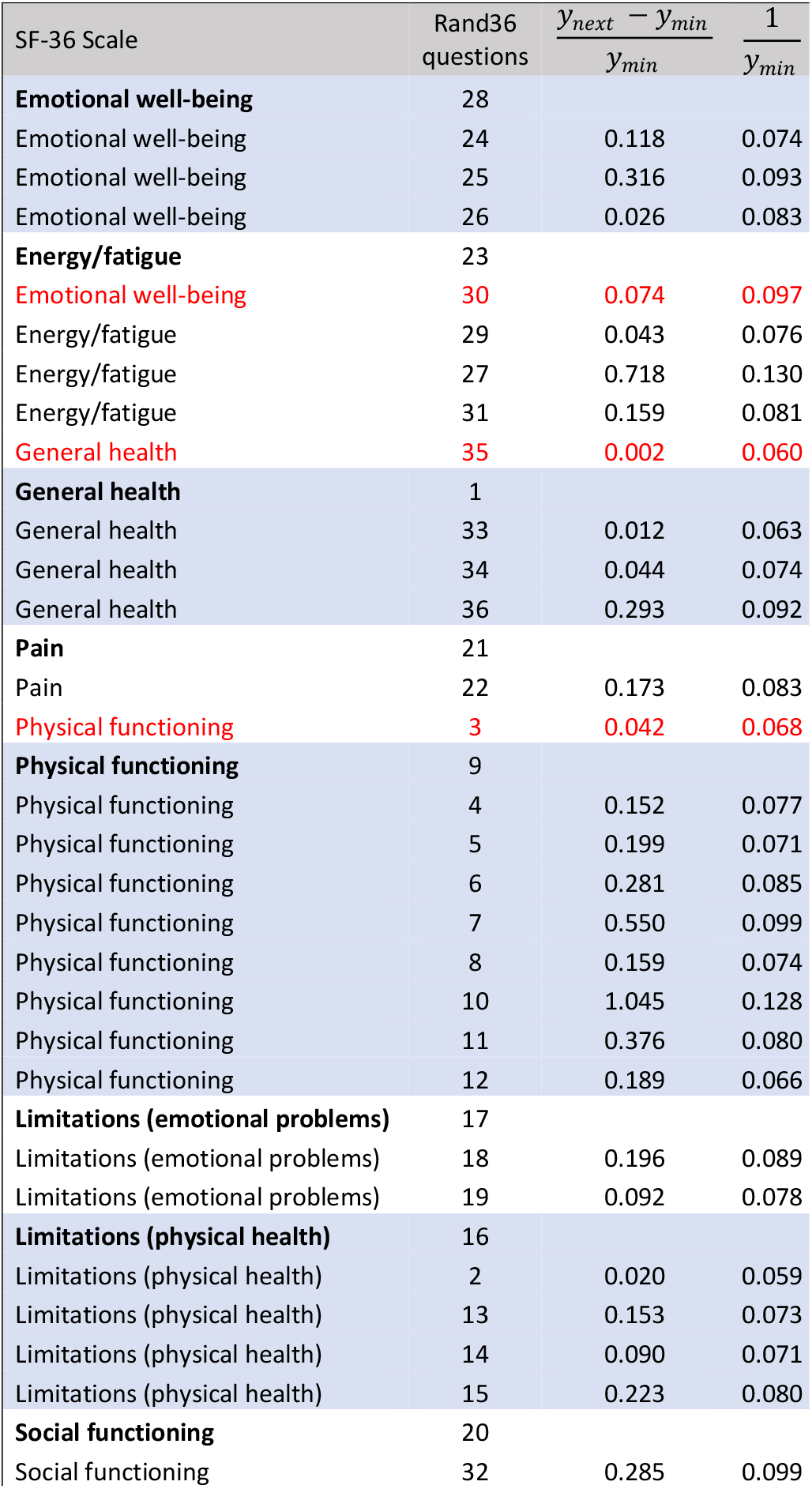
Grouping of RAND36 quality of life questions based on hierarchical clustering. The dendrogram distance between each feature (second column) and the closest landmark feature representing a health concept (first column) is denoted as *y*_*min*_, while the distance to the next closest landmark is *y*_*next*_. Each selected landmark is indicated as the first feature in the group of questions. Therefore, the fourth and third columns characterize the strength and exclusiveness of the association of each feature with its group. The dendrogram distances are measured as the Euclidean lengths between the rows of the pairwise correlation matrix. The misclassified features (Q30, Q35, and Q3) are highlighted in red.

### 2.9 Machine Learning Analysis

All computational analysis and visualizations in this work were performed using in-house scripts within the programming language and numeric computing environment MATLAB. For training machine learning models, the input data was randomly partitioned into 20% training and 80% testing sets using the ‘cvpartition’ function. Then, for the training set, an ensemble of bootstrap-aggregated (“bagged”) decision trees was built using ‘TreeBagger’ function. Bootstrap aggregation helps reduce overfitting by individual decision trees. This function selects predictors for decision splits based on the random forest algorithm [34]. To report feature importance, we used the output variable ‘OOBPermutedPredictorDeltaError.’ This variable is calculated as an increase in prediction error if the feature values are permuted between the out-of-bag observations for each tree, followed by averaging over all trees in the ensemble and dividing by the standard deviation. Finally, the predicted values for validation were calculated by applying the ‘predict’ function with the held-out test set and the ensemble of trees (i.e., the trained model) as inputs. To estimate the accuracy of the prediction, we repeated each training independently 100 times (folds) with different randomized 20/80 splits and combined the true and predicted values from all folds. Then, the aggregated true and predicted values were compared by determining the Pearson correlation coefficient and the p-value of the null hypothesis that there is no correlation (both using the ‘corrcoef’ function). The feature importance for each training procedure was calculated as an average of feature importance values from the 100 folds.

## 3. RESULTS

### 3.1 The expression of p16 is significantly different between participants grouped based on both clinical labs and the RAND36 survey

After establishing that the hierarchical clustering produces sufficiently accurate grouping of the features in the RAND36 survey (see Methods), we ask if grouping people based on their clinical labs generates subpopulations with district profiles of the other assessments (senescence biomarker, quality of life, and physical evaluation). **Figure 3** shows absolute and logarithmic values (as well as the corresponding mean and median trends) of p16 against participant age. While there is a characteristic increase in p16 with age as previously shown [35], p16 expression is highly variable between participants within each age group. Therefore, to analyze such intrinsically noisy data, we approach grouping statistically by excluding a small randomly selected subset (2 out 250) of people and repeating the hierarchical clustering 1000 times. An example of a single iteration of an unsupervised grouping of 248 people into two large clusters is shown in **Supplemental Figure 4**. The mean values and the confidence regions (±2 standard errors) for each clinical lab measurement in this example are shown in **Supplemental Figure 5A** with p-values from the two-sample t-test on the right side of the graph. Most of the clinical lab measurements are significantly different between the two clusters (p-value < 0.05). A significant difference in the clinical lab measurements is expected because these measurements were used in the clustering algorithm. However, the other (unused) measurements may or may not differ between the clusters. Indeed, **Supplemental Figure 5B** shows that only a few questions in the RAND36 survey are significantly different between the clinical lab-based subpopulations, while all outcomes of physical evaluation differ significantly (**Supplemental Figure 5C**). A plausible explanation is that physical characteristics, such as elevated blood pressure, BMI, HR, Gait, and UAG, are strongly associated with high levels of cholesterol (tot Chol, Chol/HDL-C, LDL-C, non-HDL-C), triglycerides, hematocrit, hemoglobin, glucose, HbA1c, total protein, etc., even before the onset of specific and serious health problems. On the other hand, RAND36 is designed to assess ongoing physical and mental health conditions. In other words, clinical labs and RAND36 may represent different time points of the physiological decline and, thus, do not produce fully overlapping grouping by cross-sectional data. In this regard, it is intriguing that our results (**Figure 4**) show a significant difference in p16 expression between the two groups.

**Figure 3.**
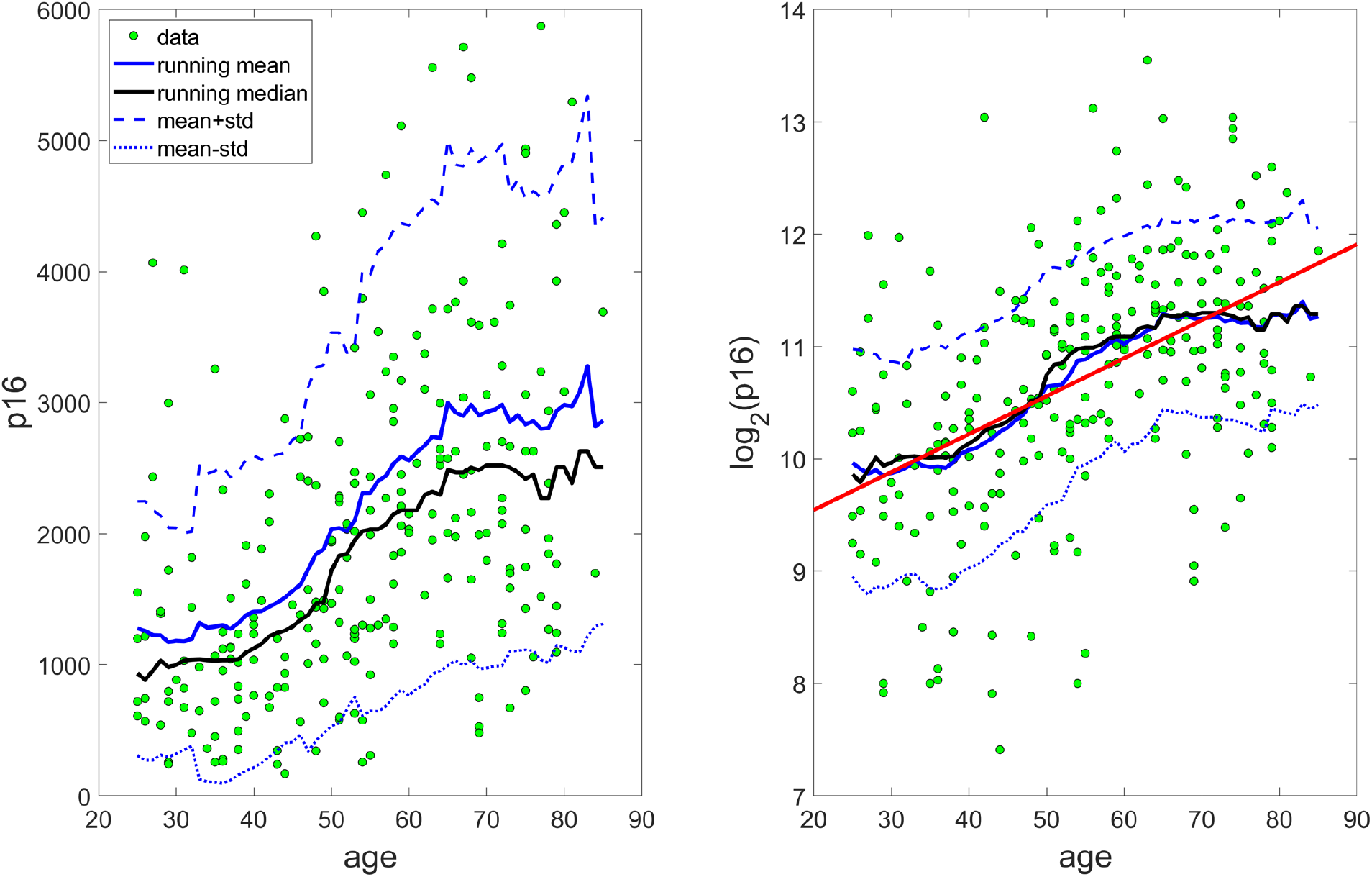
p16 expression versus age. The left and right panels show p16 expression values in linear and log^2^ scales, respectively. Solid blue lines illustrate the age trend (running average with a 20-years window size and a 1-year sliding step) with a characteristic initial phase of slow growth, followed by a fast acceleration period that ultimately levels off into a plateau. Blue dotted and dashed lines indicate the ± standard deviation range. Black lines are the running median with a 20-years window size and a 1-year sliding step. The red correlation line corresponds to *r* = 0.48 and p-value = 0.7e-15.

**Figure 4.**
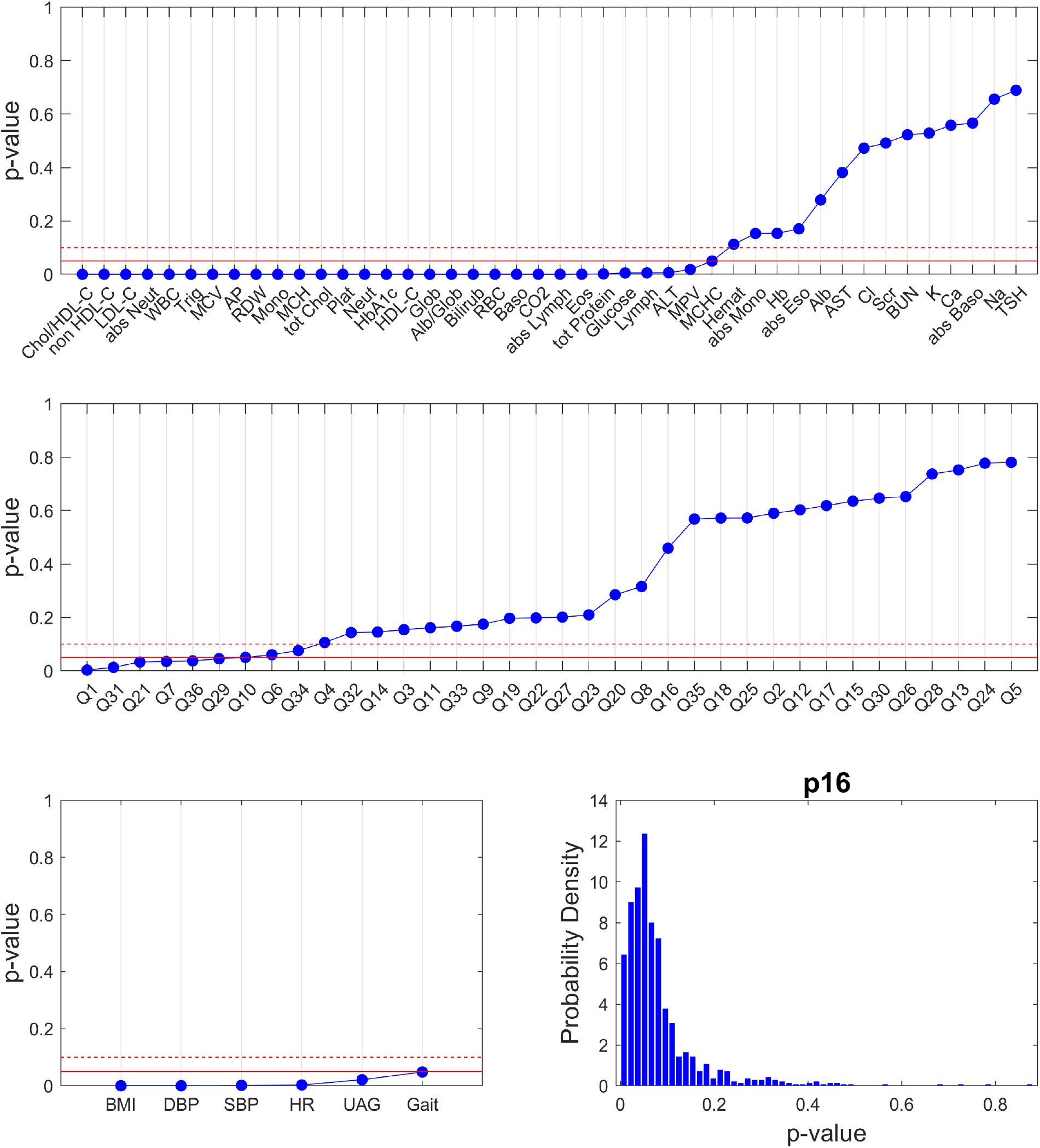
Statistical significance of differences between participants grouped based on <u>clinical lab</u> measurements using unsupervised hierarchical clustering. The clustering was performed 1000 times; each time excluding two randomly selected people and repeating the procedure for the remaining people. Here p-values validate the null hypothesis that the compared measurements in the two groups have equal mean and variance (two-sample t-test). Blue dots are the median p-values over 1000 randomized repeats for each measure in the dataset. The solid red line corresponds to p-value = 0.05 and the red dashed line to p-value = 0.1. The bottom right graph shows that distribution of p-values for p16 expressions in the resulting clinical labs-based groups.

The result for median p-values (two-sample t-test) after 1000 iterations of clinical lab-based grouping is summarized in **Figure 4**. Clinical lab measurements that are consistently different (p-value < 0.05) between the two groups (30 out of 44 measures) include the full lipid panel (6 measures), red blood cell characteristics (6 measures), most of the white blood cell types (9 measures), protein levels (tot Protein, Glob, Alb/Glob), HbA1c, Glucose, ALT, AP, and CO2. Clinical lab-based grouping shows a significant difference only for 7 out of 36 questions in the RAND36 survey. For the physical evaluation, all differences are significant. Finally, p16 varies between the two groups, with the median p-value near 0.05. Similar overall patterns were obtained for the gender- and age-corrected data (**Supplemental Figures 6 and 7**).

Similarly, we performed 1000 iterations of grouping based on RAND36 data. Now, as expected, we see a significant difference between the participant groups in the majority of the survey questions, while most of the blood measurements differ insignificantly (**Supplemental Figures 8**). The statistical results for the original (uncorrected) data are shown in **Figure 5**. Interestingly, the list of questions that differ between the groups includes all items in the role limitations due to personal or emotional problems, emotional well-being, social functioning, energy/fatigue, and general health perceptions, and about half of the physical functioning questions. The questions that differ insignificantly include items in bodily pain, role limitations due to physical health problems, and the remaining half of the physical functioning. This split can be possibly characterized as a set of questions associated with age-related issues versus a set related to age-independent (more strenuous) physical problems or damage. The senescence biomarker p16 differs significantly between the RAND36-based groups. After accounting for gender difference, p16 remains significantly different (**Supplemental Figure 9**), while the age correction leads to groups with an insignificant difference of p16 (**Supplemental Figure 10**).

**Figure 5.**
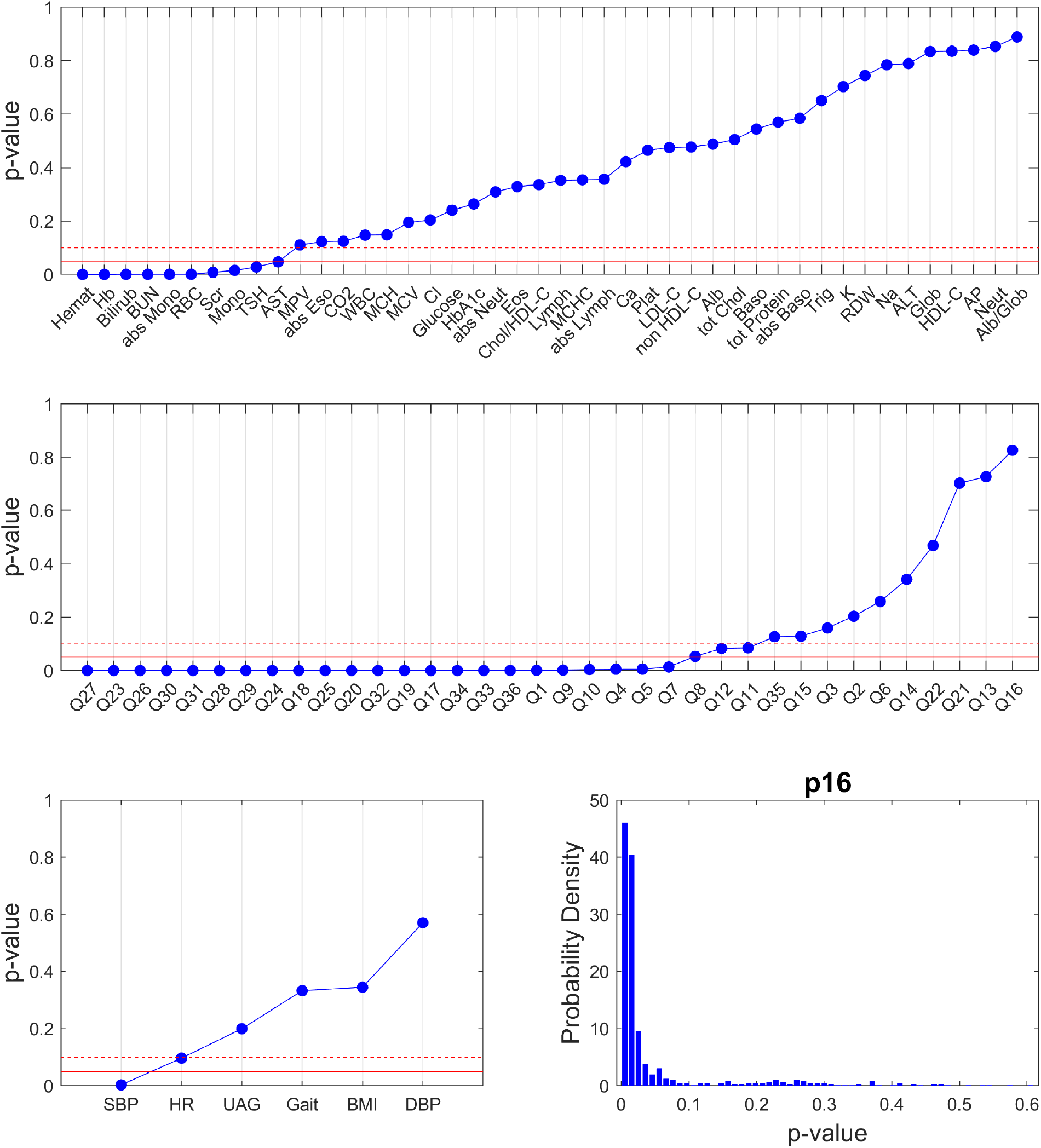
Statistical significance of differences between participants grouped based on <u>RAND36 QOL</u> responses using unsupervised hierarchical clustering. As in Figure 4, the clustering was performed 1000 times; each time excluding two randomly selected people and repeating the procedure for the remaining people. Here p-values validate the null hypothesis that the compared measurements in the two groups have equal mean and variance (two-sample t-test). Blue dots are the median p-values over 1000 randomized repeats for each measure in the dataset. The solid red line corresponds to p-value = 0.05 and the red dashed line to p-value = 0.1. The bottom right graph shows that distribution of p-values for p16 expressions in the resulting RAND36-based groups.

Overall, the major outcome of this analysis is that subpopulations with different clinical lab measurements or different health survey responses tend to have different p16. However, the clinical lab-based and RAND36-based subpopulations tend not to overlap, which suggests that p16 may serve as a biomarker that links the early (predisposing) and late (ongoing) stages of age-related health decline.

### 3.2 The highest accuracy of p16 prediction requires diverse types of measurements within their assessment categories

Predicting one of the measurements based on the others does not necessarily follow the rule “the more features, the better.” The number of features giving the most accurate prediction depends on the underlying patterns in the dataset. Here, we ask how accurately the expression of p16 can be predicted with ML models (see Methods) using 80/20 data split for training and testing. Our goal is not only to achieve the highest possible accuracy of prediction but also to determine the measures that contribute the most to the predictive power. To this end, we implement a strategy in which we start by training the model with the whole feature set, evaluate feature importance, remove the feature with the lowest importance score from the set, and repeat the process until two features are left. As a measure of accuracy, we report the correlation coefficient, *r*, between the predicted and true values of the testing dataset. Typically, this approach gives an optimal (highest accuracy) feature set between two extremes: a low accuracy for the whole set (because many features have negative importance and hurt the prediction) and the two last features (because a lot of information carried by the other features were not used for prediction). To account for the variability due to the randomization of the 80/20 split, we repeat each iteration 100 times and average the result. Finally, to account for possible variability in the feature removal sequence, we repeat the whole sequence 20 times and average the importance over all 20 repeats and over all 100 iterations (using zero importance at iterations when the feature is removed).

Figure 6. illustrates this routine for predicting p16 expression by clinical lab measurements (without gender or age correction). Here, the optimal feature set improves the accuracy from *r* = 0.19 (all 44 measures) to *r* = 0.33 (13 measures). Notably, the optimal set includes about 30% of available clinical lab measures and still represents all major types of clinical labs: diabetes screen (HbA1c), complete blood count (Hemat, Hb, RBC), blood differential (Neut, Lymph, abs Lymph, Eos, abs Eos), basic metabolic panel (BUN), comprehensive metabolic panel (AP, tot Protein), and lipid panel (tot Chol). The results for gender and age correction are shown in **Supplemental Figures 11** and **12**. Our regression-based gender normalization produces a similar result with mostly the same features at the top of the importance list but in a different order. HbA1c is still the highest importance feature. One feature that moved up to the optimal set after normalization was calcium (Ca). The optimal set for the data after age correction still makes the prediction accuracy significantly higher than the whole set (from *r* = 0.03 to *r* = 0.18), but this accuracy is about half as low as it is for the predictions with and without gender normalization. Most noticeably, after age correction, HbA1c drops from the top of the importance list to the bottom.

**Figure 6.**
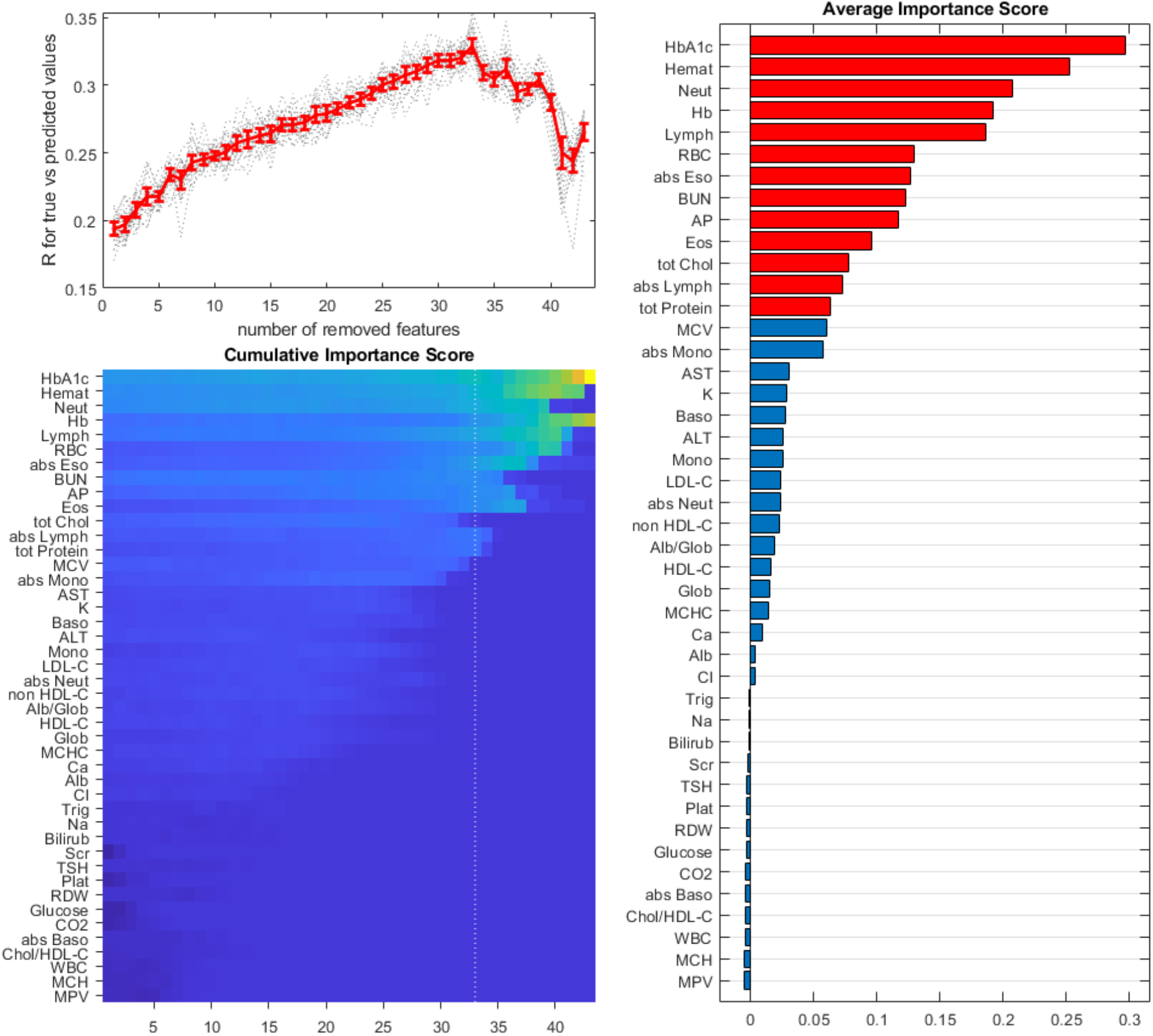
Clinical lab measurements that give the highest prediction accuracy for p16 expression in the dataset without gender or age normalization. The top left panel shows the correlation coefficient between predicted and true values in the validation set as a function of the number of iteratively removed features with the lowest importance scores. The results for 20 repeats are in gray, and the mean ± 2 standard errors are in red. The bottom left panel shows a colormap of average importance scores over 20 repeats of the feature exclusion protocol. The vertical dotted line corresponds to the exclusion step with the highest prediction accuracy. The dark blue to bright yellow colormap represents the importance scores between min to max values. The right panel shows average importance scores. The red color indicates features presented in the optimal set. The results for normalized data are shown in Supplemented Materials.

The same analysis for predicting p16 by the RAND36 data is shown in **Figure 7** (original data) and **Supplemental Figures 13** (gender normalized data) and **14** (age-corrected data). Here we have a similar situation with 12 out of 36 questions from different health concepts forming the optimal set: physical functioning (Q8, Q9, Q3, Q6), role limitations due to personal or emotional problems (Q17), emotional well-being (Q25, Q26), social functioning (Q20), energy/fatigue (Q31, Q29, Q27), and general health perceptions (Q1). Age correction gives the lowest accuracy (*r* = 0.12) and the smallest optimal set compared to the data before (*r* = 0.29) and after (*r* = 0.37) gender normalization. The results are summarized in **Table 2** and **Supplemental Table 3**.

**Figure 7.**
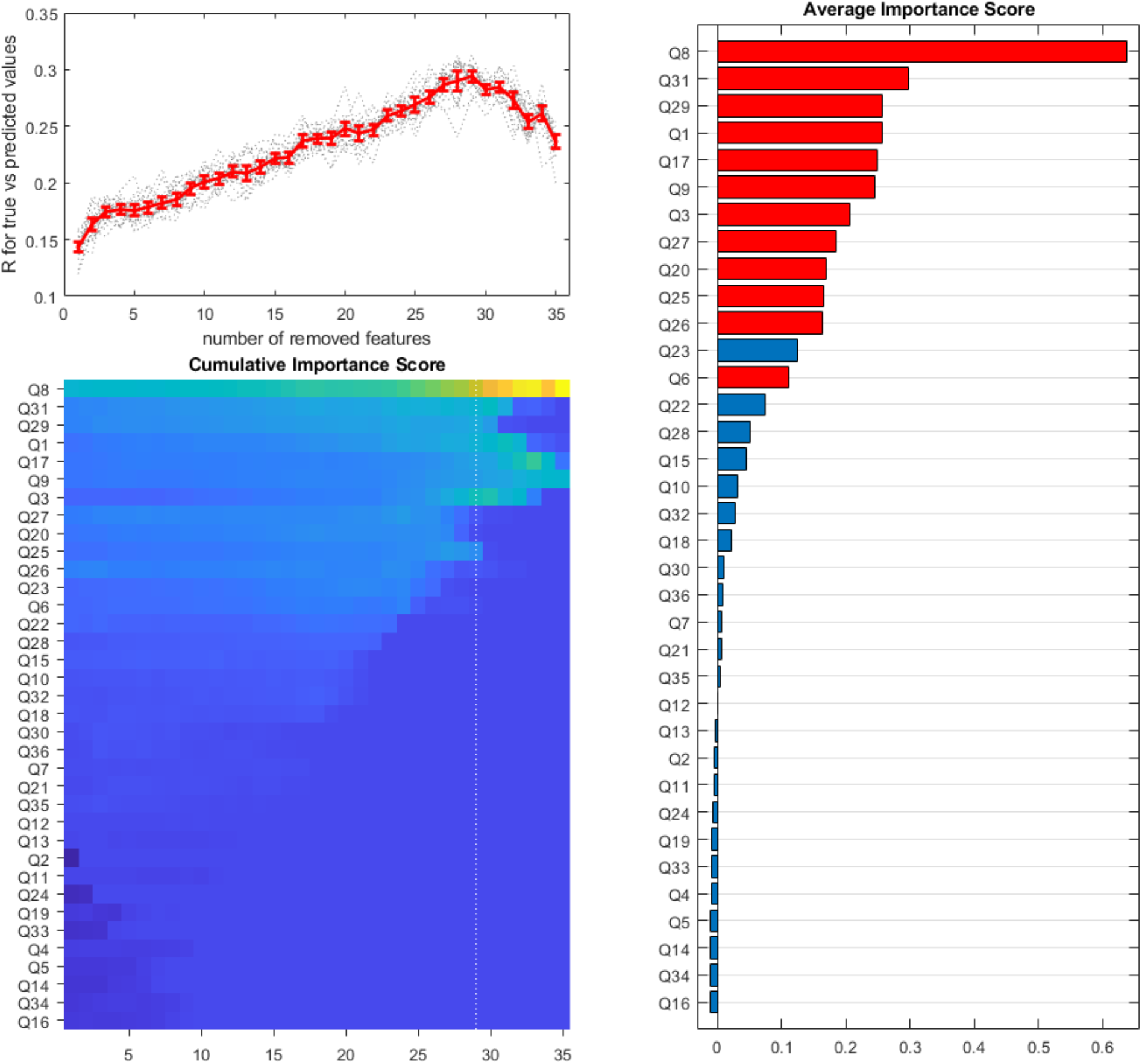
RAND36 survey responses that give the highest prediction accuracy for p16 expression in the dataset without gender or age normalization. The top left panel shows the correlation coefficient between predicted and true values in the validation set as a function of the number of iteratively removed features with the lowest importance scores. The results for 20 repeats are in gray, and the mean ± 2 standard errors are in red. The bottom left panel shows a colormap of average importance scores over 20 repeats of the feature exclusion protocol. The vertical dotted line corresponds to the exclusion step with the highest prediction accuracy. The right panel shows average importance scores. The red color indicates features presented in the optimal set. The results for normalized data are shown in Supplemented Materials.

**Table 2:**
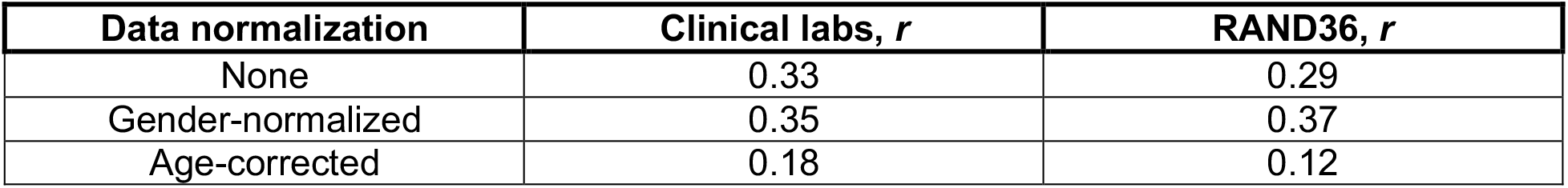
Accuracy of p16 prediction by clinical labs and RAND36 survey using 80/20 training/validation split. (reported as the average correlation coefficient between the predicted and true values)

| Data normalization | Clinical labs, $r$ | RAND36, $r$ |
| --- | --- | --- |
| None | 0.33 | 0.29 |
| Gender-normalized | 0.35 | 0.37 |
| Age-corrected | 0.18 | 0.12 |

### 3.3 Composite indexes for assessment categories provide simple overall metrics that strongly correlate with p16 expression

Training ML models to predict p16 gives a sense of how well the standard clinical tests can predict cellular senescence. However, another important question is how well senescence biomarkers can predict a person’s health condition or a predisposition for developing age-related issues. Without longitudinal data, we can’t answer this question directly, but we can use cross-sectional data to extract a single metric (index) for each assessment category based on the strength of its association with the biomarker expression and compare these indexes with the other assessments, effectively reducing the extensive feature set to a few categorical readouts. In future studies, these senescence-based indexes can be tested for a direct association with the physiological decline or development of age-related diseases. To this end, we use the following strategy. For a given assessment (*a*) and a biomarker (*b*), we define the composite index for each person (*n*) as

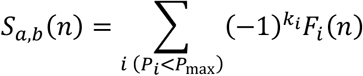

where *F*_*i*_ is the *i*’s feature (z-normalized measurement) in the assessment a; *P*_*i*_ is the p-value for the correlation between the feature *F*_*i*_ and the biomarker *b* for all people in the set; *k*_*i*_ is 0 for positive correlation and 1 for negative correlation; and the summation is performed for all features with *P*_*i*_ less than *P*_*max*_. For example, if we consider the clinical labs as an assessment, p16 as a biomarker, and *P*_*max*_ = 0.05, the 11 features in the summation are tot Chol (*P*_1_ = 0.013, *k*_1_ = 0), LDL-C (*P*_9_ = 0.044, *k*_9_ = 0), non HDL-C (*P*_3_ = 0.043, *k*_3_ = 0), BUN (*P*_4_ = 0.004, *k*_4_ = 0), K (*P*_<_ = 0.017, *k*_<_ = 0), Ca (*P*_=_ = 0.014, *k*_=_ = 0), AP (*P*_>_ = 0.033, *k*_>_ = 0), HbA1c (*P*_5_ = 0.012, *k*_5_ = 0), abs Lymph (*P*_6_ = 0.040, *k*_6_ = 1), Neut (*P*_10_ = 0.018, *k*_10_ = 0), Lymph (*P*_11_ = 0.029, *k*_11_ = 1). In this example, the index *S*_*labs*, p16_ has a correlation coefficient of 0.31 (p-value 5.8e-7) with p16, which is close to the optimal prediction accuracy of 0.33 using our ML training approach (see **Figure 6**). The fact that a simple linear combination produces a single clinical lab-based index so strongly correlating with the biomarker of interest justifies our scoring approach, which doesn’t require complex algorithms and extensive model training.

To illuminate the need to choose a specific cutoff for p-value, *P*_*max*_, we calculated the index for the full range of values between 0.01 and 1 with a 0.01 step. **Figure 8** shows the coefficient of correlation between the index and p16 and the ratio of this value to the *maximal* coefficient of correlation between p16 and the individual features in the assessment set. The latter characteristic tells us how much the composite index improves correlation over a single most strongly correlating feature. **Supplemental Table 4** lists the measures for clinical lab-based and RAND36-based indexes that give the largest improvement for the correlation with p16.

**Figure 8.**
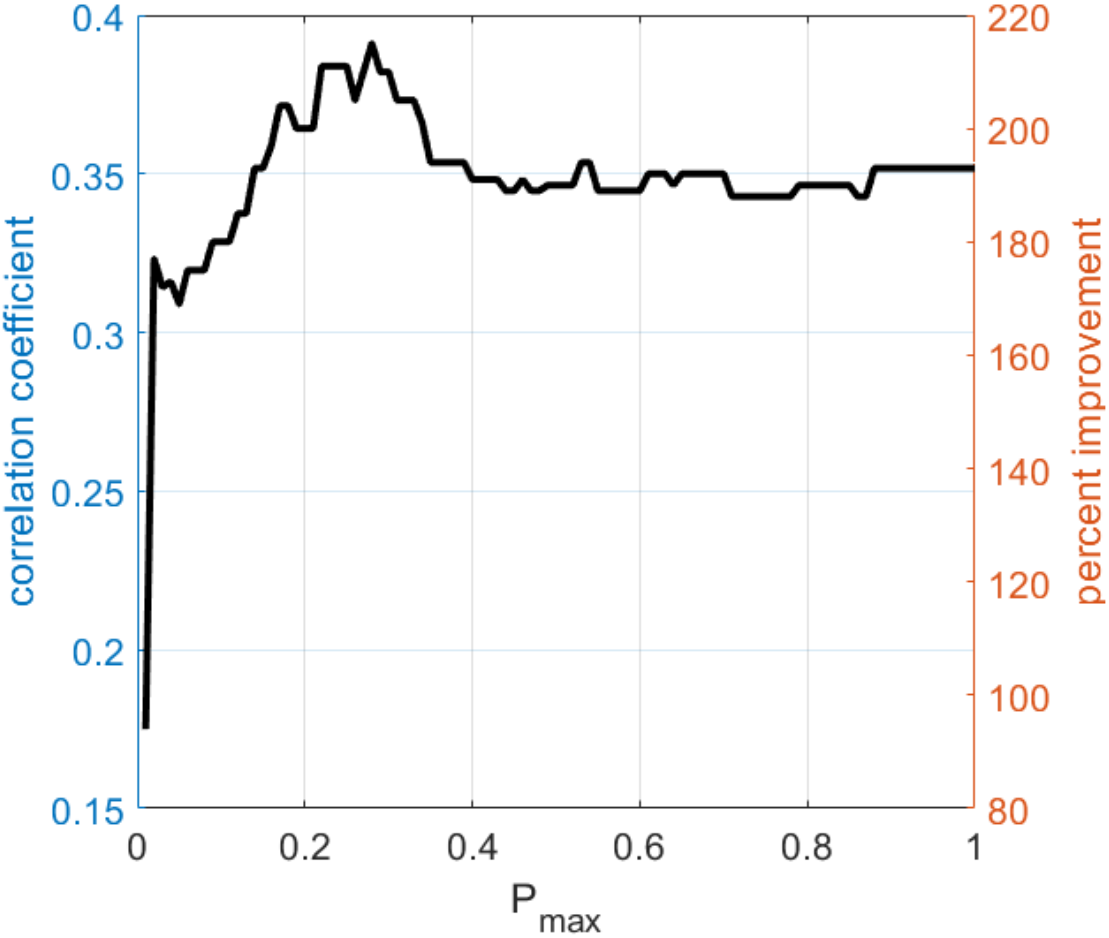
An illustration of the linear combination approach to finding the score metric with the highest correlation with p16. For each value of the cutoff, P_max_, clinical lab features with the higher individual correlation significances (p-value<P_max_) are combined in a single multi-feature measure. The left axis indicates the correlation coefficient between the resulting score and the p16 expression. The right axis shows the ratio of this coefficient to the highest correlation coefficient of a single feature in the set, indicating the correlation improvement, so that P_max_ value corresponding to the highest improvement indicates the best combination of measurements.

Once again, we see that p16 strongly correlates with both clinical labs and the RAND36 survey. The correlation between assessment indexes *S*_*labs*, p16_ and *S*_rand36, p16_ is significant but not as strong (*r* < 0.3) as their correlation with p16. We can propose two potential explanations. One is that clinical lab-based and RAND36-based indexes represent sufficiently different aspects of age-related health decline, but since p16 is indicative of system-level aging, it correlates strongly with each of the assessments. Another possibility, and the one that we are leaning towards, is that clinical lab-based and RAND36-based indexes represent earlier and later phases of decline. In this case, a high value for *S*_*labs*, p16_, is a predictor of a trajectory towards developing a high value for *S*_rand, p16_ for the same individuals later in their lives. Confirming the validity of the second interpretation requires a longitudinal study of the senescence biomarkers. The composite indexes developed here (**Supplemental Table 4**) can help to simplify such temporal tracking.

## 4 DISCUSSION

In this report, we analyzed interrelationships between the expression of a gene used to measure cellular senescence, p16, and three batteries of assessments: clinical labs (44 measures), RAND36 quality-of-life survey (36 measures), and physical evaluation (6 measures). We approached the analysis several ways. First, we checked to what degree pairwise correlations between measurements are mediated by their common correlation with age. To this end, we compared the data adjusted for the gender differences before and after additional correction for the age trend. In this analysis, the p16 expression was strongly affected by the age correction. Only five measurements (total protein, absolute eosinophil count, absolute and relative basophil count, and question 35, “I expect my health to get worse”) have a significant correlation with p16 when the age trends are subtracted (comparing only the deviations from the trend). That means, for example, that people with a p16 level elevated with respect to the mean level for their age are more likely to have an increased level of serum total protein and lower absolute counts of eosinophils and basophils (as compared to the mean values for their age group). Two measurements (potassium and calcium) have strong positive correlation with p16 regardless of age correction.

Next, we determined how grouping people based on one assessment reflects the differences between the resulting groups in terms of the other assessments. For classification and comparison, we only considered the splits into two major groups using unsupervised hierarchical clustering. Before using this method, we verified that our data had sufficient statistical power to reproduce a previously established grouping of RAND36 QOL questions into eight health concepts. Our hierarchical clustering misclassified only three questions at the borderline between pairs of concepts. Application of this method to group people has shown that clinical lab-based and RAND36-based groupings produce weakly overlapping subpopulations. This distinction between the two assessments may reflect the fact that deviation of a clinical lab measurements from a norm is indicative of a developing issue (e.g., elevated HbA1c vs diagnosed diabetes), while the self-assessment reports an ongoing physical or mental limitation (e.g., difficulty in lifting or carrying groceries). Thus, if we interpret clinical labs and RAND36 assessments as indicators of earlier and later phases of physiological decline, it is not surprising that these assessments classify people in a mixed population differently. The clinical lab-based clustering produced groups with significantly different outcomes of physical evaluation (all six measures), while RAND36-based groups only differed in the systolic blood pressure. An intriguing result is that p16 turned out to be significantly different in the groups segregated by clinical labs and RAND36, while there was no difference in the average age of the people in these groups. Thus, p16 and not participants’ age is indicative of ongoing health limitations but also sensitive to developing issues before the onset of a serious condition. In other words, p16 may serve as an earlier predictor of the physiological decline. This suggestion needs to be verified by a longitudinal study tracking p16 expression and health state over the course of time.

Following the unsupervised classification, we sought to explore the accuracy of predicting biomarker expressions based on clinical labs and RAND36 survey using the standard machine learning approach: training the model on 80% of the data and verifying the predicted values for the held out 20% of the data. We determined that optimal subsets of measurements providing the highest accuracy (i.e., the largest correlation coefficient between the true and predicted values) are typically about two-fold larger than the strongest correlation coefficient between p16 and the individual measurements in the assessments. Interestingly, such optimal feature subsets for p16 tend to include diverse features covering clinical lab measurements from different panels or RAND36 questions from different health concepts. We interpret this result as evidence that p16, a biomarker of accumulating cellular senescence, is also a biomarker of the system-level physiological state (i.e., the biological age).

Because many of the measures contributing to the accuracy of the predictions have statistically significant (p-value < 0.05, *r*∼0.15) correlations with p16, we tested if a simple addition of measures (with positive and negative signs for positively and negatively correlating measures, respectively) can give us metrics that strongly correlate with the biomarker. Indeed, this approach produced composite indexes (combined subsets of measurements from each assessment) with correlation coefficients equivalent to the correlations between true and predicted values (*r*∼0.35) by the machine learning approach. Obviously, such an algorithm is not a better option or a substitute for machine learning methods in general. A larger dataset could provide significantly improved training accuracy and capture more complex interrelations in the data than a simple linear combination. However, for our 250 (people) x 87 (measures) dataset, the linear indexes give fast, simple, and interpretable metrics correlating with p16 significantly (over twofold) stronger than the individual measurements. This way, we reduced multivariable assessments to a characterization with only two indexes representing the different levels of cellular senescence in individuals.

Previously, we developed a two-component mathematical model of p16 accumulation with age that accurately reproduced both the mean trends and the variability of the gene expression as a function of time [35]. This mechanistic model described p16 changes over time, with the characteristic exponential growth early and saturation later in life, because of an interplay between the accumulation and clearance of senescent cells. If we interpret the rate of clearance as the efficiency of the immune system (or the level of immune senescence), the model predicts that the highest level of senescent cells will result from simultaneously high rates of cellular and immune senescence; the lowest level from simultaneously low rates, and the intermediate levels when only one of the rates is elevated. The new data presented here suggests that p16 alone is a strong indicator of physiological differences at the earlier and later stages of the health decline. However, consolidating such differences across different stages would require a combination of both cellular and immune senescence biomarkers.

## Data Availability

All data produced in the present study are available upon reasonable request to the authors

## AUTHOR CONTRIBUTIONS

L. T. and D. T. performed computational analysis, prepared figures, and drafted the manuscript. S. H. performed initial data processing and early stages of computational analysis. A. R. recruited participants for the study and curated data. K. N. and H. M. oversaw participant recruitment and data collection. N. M. supervised senescence gene expression analyses. L. T., A. R., and D. T. developed computational methodology. L. T., K. N., H. M., N. M., A.E., and D. T. edited the manuscript. H. M., N. M., and D. T. conceived the study and secured funding for the project. D. T. supervised the overall project direction. All authors contributed to the manuscript discussion and provided feedback that helped shape the research process.

## SUPPLEMENTAL MATERIALS

**Supplemental Figure 1.**
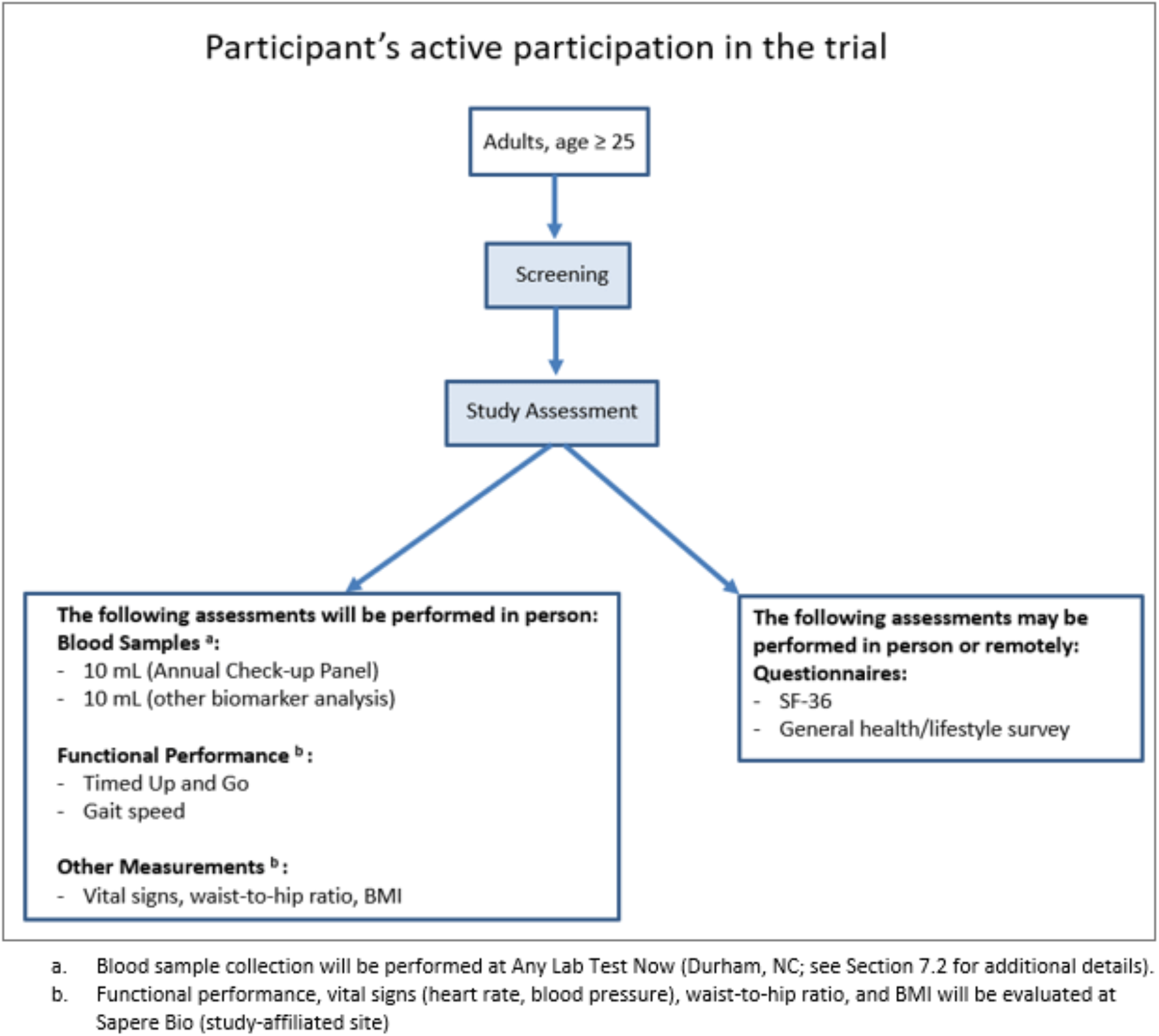
Study Schematic.

**Supplemental Table 1:** Statistical gender differences for clinical labs before and after normalization. Colors highlight p-values <0.05 (pink), >0.10 (green), and >0.05 & <0.1 (yellow).

|  | not normalized |  |  |  |  |  |  |  |  |  |  | normalized |  |  |  |  |  |  |
| --- | --- | --- | --- | --- | --- | --- | --- | --- | --- | --- | --- | --- | --- | --- | --- | --- | --- | --- |
| | female | | | | | male | | | | | $p_{MF}$ | female | | | male | | | $p_{MF}$ |
| measure | mean | std | se | r | $p_{corr}$ | mean | std | se | r | $p_{corr}$ | | mean | std | se | mean | std | se | |
| p16 | 10.70 | 1.16 | 0.09 | 0.48 | 0.00 | 10.72 | 0.91 | 0.11 | 0.51 | 0.00 | 0.86 | 10.70 | 1.16 | 0.09 | 10.91 | 0.98 | 0.12 | 0.17 |
| tot Chol | 194.69 | 34.39 | 2.56 | 0.31 | 0.00 | 184.32 | 39.41 | 4.74 | 0.08 | 0.52 | 0.04 | 194.69 | 34.39 | 2.56 | 198.77 | 40.80 | 4.91 | 0.43 |
| HDL-C | 69.01 | 14.49 | 1.08 | 0.22 | 0.00 | 56.25 | 14.29 | 1.72 | -0.07 | 0.59 | 0.00 | 69.01 | 14.49 | 1.08 | 70.23 | 14.63 | 1.76 | 0.55 |
| Trig | 90.25 | 43.25 | 3.21 | 0.13 | 0.07 | 110.46 | 79.07 | 9.52 | 0.13 | 0.27 | 0.01 | 90.25 | 43.25 | 3.21 | 92.46 | 78.58 | 9.46 | 0.78 |
| LDL-C | 107.01 | 30.99 | 2.30 | 0.21 | 0.00 | 107.44 | 36.19 | 4.39 | 0.09 | 0.45 | 0.93 | 107.01 | 30.99 | 2.30 | 109.41 | 36.66 | 4.45 | 0.60 |
| Chol/HDL-C | 2.92 | 0.77 | 0.06 | 0.06 | 0.46 | 3.47 | 1.24 | 0.15 | 0.10 | 0.40 | 0.00 | 2.92 | 0.77 | 0.06 | 2.94 | 1.23 | 0.15 | 0.90 |
| non HDL-C | 125.66 | 34.19 | 2.54 | 0.22 | 0.00 | 128.07 | 40.09 | 4.83 | 0.10 | 0.41 | 0.64 | 125.66 | 34.19 | 2.54 | 128.52 | 40.63 | 4.89 | 0.58 |
| Glucose | 90.71 | 14.00 | 1.04 | 0.22 | 0.00 | 96.20 | 33.55 | 4.04 | 0.14 | 0.26 | 0.07 | 90.71 | 14.00 | 1.04 | 91.87 | 33.38 | 4.02 | 0.70 |
| BUN | 13.97 | 3.93 | 0.29 | 0.51 | 0.00 | 16.45 | 5.20 | 0.63 | 0.38 | 0.00 | 0.00 | 13.97 | 3.93 | 0.29 | 14.73 | 5.23 | 0.63 | 0.21 |
| Scr | 0.77 | 0.12 | 0.01 | 0.15 | 0.05 | 1.00 | 0.27 | 0.03 | 0.17 | 0.16 | 0.00 | 0.77 | 0.12 | 0.01 | 0.77 | 0.26 | 0.03 | 0.78 |
| Na | 139.28 | 2.00 | 0.15 | 0.15 | 0.05 | 139.49 | 1.88 | 0.23 | -0.16 | 0.18 | 0.45 | 139.28 | 2.00 | 0.15 | 139.39 | 1.88 | 0.23 | 0.69 |
| K | 4.40 | 0.32 | 0.02 | 0.06 | 0.41 | 4.48 | 0.33 | 0.04 | 0.09 | 0.46 | 0.10 | 4.40 | 0.32 | 0.02 | 4.41 | 0.33 | 0.04 | 0.87 |
| Cl | 103.94 | 2.09 | 0.16 | -0.11 | 0.12 | 103.77 | 2.07 | 0.25 | 0.06 | 0.62 | 0.56 | 103.94 | 2.09 | 0.16 | 103.85 | 2.08 | 0.25 | 0.76 |
| CO2 | 25.93 | 1.99 | 0.15 | 0.24 | 0.00 | 26.46 | 2.13 | 0.26 | -0.21 | 0.08 | 0.06 | 25.93 | 1.99 | 0.15 | 26.11 | 2.14 | 0.26 | 0.53 |
| Ca | 9.50 | 0.36 | 0.03 | 0.11 | 0.16 | 9.52 | 0.33 | 0.04 | -0.19 | 0.11 | 0.63 | 9.50 | 0.36 | 0.03 | 9.51 | 0.32 | 0.04 | 0.77 |
| tot Protein | 6.88 | 0.35 | 0.03 | -0.17 | 0.02 | 6.77 | 0.37 | 0.04 | -0.16 | 0.19 | 0.03 | 6.88 | 0.35 | 0.03 | 6.86 | 0.37 | 0.04 | 0.64 |
| Alb | 4.38 | 0.23 | 0.02 | -0.14 | 0.07 | 4.39 | 0.26 | 0.03 | -0.39 | 0.00 | 0.85 | 4.38 | 0.23 | 0.02 | 4.37 | 0.24 | 0.03 | 0.72 |
| Glob | 2.50 | 0.30 | 0.02 | -0.10 | 0.17 | 2.38 | 0.30 | 0.04 | 0.15 | 0.23 | 0.00 | 2.50 | 0.30 | 0.02 | 2.49 | 0.30 | 0.04 | 0.78 |
| Alb/Glob | 1.78 | 0.24 | 0.02 | 0.03 | 0.69 | 1.87 | 0.26 | 0.03 | -0.29 | 0.02 | 0.01 | 1.78 | 0.24 | 0.02 | 1.78 | 0.25 | 0.03 | 0.94 |
| Bilirub | 0.61 | 0.22 | 0.02 | 0.10 | 0.17 | 0.85 | 0.31 | 0.04 | 0.00 | 0.99 | 0.00 | 0.61 | 0.22 | 0.02 | 0.62 | 0.31 | 0.04 | 0.80 |
| AP | 62.52 | 18.80 | 1.40 | 0.28 | 0.00 | 62.12 | 18.98 | 2.28 | 0.12 | 0.31 | 0.88 | 62.52 | 18.80 | 1.40 | 64.51 | 19.58 | 2.36 | 0.46 |
| AST | 19.09 | 5.96 | 0.44 | 0.08 | 0.29 | 22.16 | 6.72 | 0.81 | -0.18 | 0.14 | 0.00 | 19.09 | 5.96 | 0.44 | 19.27 | 6.62 | 0.80 | 0.84 |
| ALT | 16.56 | 8.86 | 0.66 | -0.01 | 0.91 | 22.65 | 10.97 | 1.32 | -0.19 | 0.11 | 0.00 | 16.56 | 8.86 | 0.66 | 16.53 | 10.76 | 1.30 | 0.98 |
| HbA1c | 5.34 | 0.46 | 0.03 | 0.32 | 0.00 | 5.38 | 0.54 | 0.06 | 0.22 | 0.07 | 0.59 | 5.34 | 0.46 | 0.03 | 5.40 | 0.55 | 0.07 | 0.41 |
| TSH | 2.08 | 1.03 | 0.08 | 0.23 | 0.00 | 2.44 | 1.53 | 0.18 | 0.07 | 0.55 | 0.04 | 2.08 | 1.03 | 0.08 | 2.17 | 1.54 | 0.19 | 0.60 |
| WBC | 5.48 | 1.44 | 0.11 | -0.03 | 0.66 | 5.70 | 1.83 | 0.22 | 0.14 | 0.25 | 0.32 | 5.48 | 1.44 | 0.11 | 5.46 | 1.82 | 0.22 | 0.93 |
| RBC | 4.48 | 0.36 | 0.03 | 0.00 | 0.98 | 4.93 | 0.38 | 0.05 | -0.22 | 0.07 | 0.00 | 4.48 | 0.36 | 0.03 | 4.48 | 0.37 | 0.04 | 0.99 |
| Hb | 13.46 | 1.03 | 0.08 | 0.09 | 0.21 | 15.22 | 1.02 | 0.12 | 0.02 | 0.87 | 0.00 | 13.46 | 1.03 | 0.08 | 13.50 | 1.02 | 0.12 | 0.80 |
| Hemat | 40.74 | 2.80 | 0.21 | 0.12 | 0.10 | 45.36 | 2.95 | 0.36 | 0.05 | 0.67 | 0.00 | 40.74 | 2.80 | 0.21 | 40.87 | 2.97 | 0.36 | 0.75 |
| MCV | 91.06 | 4.72 | 0.35 | 0.17 | 0.02 | 92.08 | 3.90 | 0.47 | 0.50 | 0.00 | 0.11 | 91.06 | 4.72 | 0.35 | 91.36 | 3.48 | 0.42 | 0.62 |
| MCH | 30.10 | 1.97 | 0.15 | 0.12 | 0.12 | 30.91 | 1.53 | 0.18 | 0.39 | 0.00 | 0.00 | 30.10 | 1.97 | 0.15 | 30.18 | 1.43 | 0.17 | 0.73 |
| MCHC | 33.04 | 0.96 | 0.07 | -0.04 | 0.63 | 33.56 | 0.81 | 0.10 | -0.09 | 0.48 | 0.00 | 33.04 | 0.96 | 0.07 | 33.02 | 0.80 | 0.10 | 0.92 |
| RDW | 12.65 | 0.75 | 0.06 | 0.08 | 0.27 | 12.57 | 0.63 | 0.08 | -0.07 | 0.54 | 0.43 | 12.65 | 0.75 | 0.06 | 12.68 | 0.64 | 0.08 | 0.82 |
| Plat | 263.29 | 59.22 | 4.40 | 0.00 | 0.95 | 233.52 | 53.40 | 6.43 | -0.16 | 0.18 | 0.00 | 263.29 | 59.22 | 4.40 | 263.40 | 52.69 | 6.34 | 0.99 |
| MPV | 10.51 | 0.95 | 0.07 | -0.14 | 0.06 | 10.53 | 0.91 | 0.11 | -0.28 | 0.02 | 0.87 | 10.51 | 0.95 | 0.07 | 10.46 | 0.89 | 0.11 | 0.70 |
| abs Neut | 3172.40 | 1094.68 | 81.37 | 0.01 | 0.86 | 3192.84 | 1297.01 | 156.14 | 0.22 | 0.08 | 0.90 | 3172.40 | 1094.68 | 81.37 | 3177.95 | 1266.56 | 152.48 | 0.97 |
| abs Lymph | 1703.88 | 505.94 | 37.61 | -0.14 | 0.05 | 1765.14 | 562.21 | 67.68 | -0.06 | 0.61 | 0.41 | 1703.88 | 505.94 | 37.61 | 1676.09 | 566.07 | 68.15 | 0.71 |
| abs Mono | 424.84 | 109.94 | 8.17 | 0.03 | 0.70 | 519.20 | 180.77 | 21.76 | 0.09 | 0.45 | 0.00 | 424.84 | 109.94 | 8.17 | 426.03 | 180.03 | 21.67 | 0.95 |
| abs Eso | 138.08 | 87.87 | 6.53 | 0.05 | 0.48 | 180.33 | 137.08 | 16.50 | -0.04 | 0.72 | 0.00 | 138.08 | 87.87 | 6.53 | 139.84 | 137.03 | 16.50 | 0.90 |
| abs Baso | 44.01 | 17.45 | 1.30 | 0.14 | 0.06 | 44.01 | 19.21 | 2.31 | 0.10 | 0.43 | 1.00 | 44.01 | 17.45 | 1.30 | 44.94 | 19.28 | 2.32 | 0.72 |
| Neut | 57.02 | 8.14 | 0.60 | 0.08 | 0.30 | 55.21 | 8.19 | 0.99 | 0.38 | 0.00 | 0.12 | 57.02 | 8.14 | 0.60 | 57.26 | 7.59 | 0.91 | 0.83 |
| Lymph | 31.64 | 7.23 | 0.54 | -0.14 | 0.06 | 31.45 | 6.37 | 0.77 | -0.38 | 0.00 | 0.85 | 31.64 | 7.23 | 0.54 | 31.25 | 5.98 | 0.72 | 0.69 |
| Mono | 7.93 | 1.78 | 0.13 | 0.11 | 0.13 | 9.27 | 2.17 | 0.26 | -0.15 | 0.23 | 0.00 | 7.93 | 1.78 | 0.13 | 8.00 | 2.16 | 0.26 | 0.77 |
| Eos | 2.59 | 1.63 | 0.12 | 0.08 | 0.30 | 3.29 | 2.52 | 0.30 | -0.14 | 0.24 | 0.01 | 2.59 | 1.63 | 0.12 | 2.64 | 2.50 | 0.30 | 0.86 |
| Baso | 0.82 | 0.31 | 0.02 | 0.17 | 0.03 | 0.80 | 0.31 | 0.04 | -0.06 | 0.65 | 0.57 | 0.82 | 0.31 | 0.02 | 0.84 | 0.32 | 0.04 | 0.65 |

**Supplemental Table 2:** Statistical gender differences for RAND36 survey and physical evaluation before and after normalization. Colors highlight p-values <0.05 (pink), >0.10 (green), and >0.05 & <0.1 (yellow).

| measure | not normalized |  |  |  |  |  |  |  |  |  |  | normalized |  |  |  |  |  |  |
| --- | --- | --- | --- | --- | --- | --- | --- | --- | --- | --- | --- | --- | --- | --- | --- | --- | --- | --- |
| | female | | | | | male | | | | | $p_{MF}$ | female | | | male | | | $p_{MF}$ |
| | mean | std | se | r | $p_{corr}$ | mean | std | se | r | $p_{corr}$ | | mean | std | se | mean | std | se | |
| Q1 | 74.03 | 19.30 | 1.44 | 0.00 | 0.97 | 78.26 | 18.15 | 2.18 | -0.01 | 0.92 | 0.12 | 74.03 | 19.30 | 1.44 | 74.00 | 18.14 | 2.18 | 0.99 |
| Q2 | 55.28 | 19.60 | 1.46 | 0.05 | 0.47 | 53.68 | 18.96 | 2.30 | -0.02 | 0.86 | 0.56 | 55.28 | 19.60 | 1.46 | 55.67 | 18.99 | 2.30 | 0.89 |
| Q3 | 64.61 | 35.02 | 2.62 | -0.32 | 0.00 | 63.97 | 33.28 | 4.04 | -0.36 | 0.00 | 0.90 | 64.61 | 35.02 | 2.62 | 60.18 | 33.20 | 4.03 | 0.37 |
| Q4 | 93.89 | 19.53 | 1.46 | -0.30 | 0.00 | 93.48 | 16.96 | 2.04 | -0.09 | 0.49 | 0.88 | 93.89 | 19.53 | 1.46 | 91.66 | 17.91 | 2.16 | 0.41 |
| Q5 | 96.94 | 12.01 | 0.90 | -0.16 | 0.03 | 96.27 | 13.24 | 1.62 | -0.03 | 0.79 | 0.70 | 96.94 | 12.01 | 0.90 | 96.21 | 13.38 | 1.63 | 0.68 |
| Q6 | 83.43 | 28.48 | 2.13 | -0.28 | 0.00 | 91.30 | 20.93 | 2.52 | -0.09 | 0.46 | 0.04 | 83.43 | 28.48 | 2.13 | 80.28 | 22.37 | 2.69 | 0.41 |
| Q7 | 96.11 | 14.43 | 1.08 | -0.17 | 0.02 | 97.10 | 11.77 | 1.42 | -0.13 | 0.29 | 0.61 | 96.11 | 14.43 | 1.08 | 95.16 | 11.94 | 1.44 | 0.63 |
| Q8 | 85.75 | 26.62 | 1.99 | -0.26 | 0.00 | 82.61 | 29.49 | 3.55 | -0.30 | 0.01 | 0.42 | 85.75 | 26.62 | 1.99 | 82.96 | 29.04 | 3.50 | 0.47 |
| Q9 | 91.06 | 25.49 | 1.91 | -0.25 | 0.00 | 88.97 | 27.11 | 3.29 | -0.18 | 0.14 | 0.57 | 91.06 | 25.49 | 1.91 | 88.54 | 27.45 | 3.33 | 0.50 |
| Q10 | 94.97 | 17.65 | 1.32 | -0.18 | 0.01 | 93.48 | 20.85 | 2.51 | -0.11 | 0.38 | 0.57 | 94.97 | 17.65 | 1.32 | 93.75 | 20.99 | 2.53 | 0.64 |
| Q11 | 98.31 | 10.52 | 0.79 | -0.05 | 0.47 | 98.53 | 8.51 | 1.03 | -0.17 | 0.18 | 0.88 | 98.31 | 10.52 | 0.79 | 98.10 | 8.41 | 1.02 | 0.88 |
| Q12 | 99.16 | 8.36 | 0.63 | -0.03 | 0.67 | 98.53 | 8.51 | 1.03 | -0.04 | 0.73 | 0.60 | 99.16 | 8.36 | 0.63 | 99.05 | 8.51 | 1.03 | 0.93 |
| Q13 | 93.89 | 24.02 | 1.79 | -0.16 | 0.03 | 92.75 | 26.12 | 3.14 | 0.06 | 0.65 | 0.74 | 93.89 | 24.02 | 1.79 | 92.38 | 26.38 | 3.18 | 0.67 |
| Q14 | 84.44 | 36.34 | 2.71 | -0.11 | 0.12 | 83.82 | 37.10 | 4.50 | -0.18 | 0.13 | 0.91 | 84.44 | 36.34 | 2.71 | 82.89 | 36.71 | 4.45 | 0.76 |
| Q15 | 91.01 | 28.68 | 2.15 | -0.27 | 0.00 | 81.16 | 39.39 | 4.74 | -0.20 | 0.11 | 0.03 | 91.01 | 28.68 | 2.15 | 88.03 | 39.44 | 4.75 | 0.51 |
| Q16 | 90.56 | 29.33 | 2.19 | -0.13 | 0.08 | 86.96 | 33.92 | 4.08 | -0.09 | 0.46 | 0.41 | 90.56 | 29.33 | 2.19 | 89.08 | 34.01 | 4.09 | 0.74 |
| Q17 | 81.56 | 38.89 | 2.91 | 0.11 | 0.15 | 89.86 | 30.41 | 3.66 | 0.02 | 0.88 | 0.11 | 81.56 | 38.89 | 2.91 | 83.21 | 30.72 | 3.70 | 0.75 |
| Q18 | 68.89 | 46.42 | 3.46 | 0.16 | 0.03 | 82.35 | 38.41 | 4.66 | -0.03 | 0.80 | 0.03 | 68.89 | 46.42 | 3.46 | 72.07 | 39.14 | 4.75 | 0.62 |
| Q19 | 83.33 | 37.37 | 2.79 | 0.19 | 0.01 | 94.20 | 23.54 | 2.83 | 0.24 | 0.04 | 0.03 | 83.33 | 37.37 | 2.79 | 86.09 | 23.99 | 2.89 | 0.57 |
| Q20 | 87.99 | 18.23 | 1.36 | 0.11 | 0.16 | 91.67 | 14.64 | 1.76 | 0.27 | 0.03 | 0.14 | 87.99 | 18.23 | 1.36 | 88.73 | 14.25 | 1.72 | 0.76 |
| Q21 | 74.81 | 20.26 | 1.51 | -0.15 | 0.05 | 71.01 | 20.73 | 2.50 | -0.11 | 0.35 | 0.19 | 74.81 | 20.26 | 1.51 | 73.67 | 20.82 | 2.51 | 0.70 |
| Q22 | 89.77 | 16.53 | 1.25 | -0.12 | 0.11 | 88.77 | 16.90 | 2.03 | -0.09 | 0.48 | 0.67 | 89.77 | 16.53 | 1.25 | 88.94 | 16.96 | 2.04 | 0.72 |
| Q23 | 57.11 | 23.84 | 1.78 | 0.22 | 0.00 | 65.80 | 20.61 | 2.48 | 0.08 | 0.51 | 0.01 | 57.11 | 23.84 | 1.78 | 59.14 | 21.24 | 2.56 | 0.54 |
| Q24 | 75.98 | 20.81 | 1.56 | 0.21 | 0.00 | 85.29 | 19.12 | 2.32 | 0.19 | 0.11 | 0.00 | 75.98 | 20.81 | 1.56 | 77.77 | 19.29 | 2.34 | 0.54 |
| Q25 | 91.16 | 14.35 | 1.07 | 0.01 | 0.90 | 95.36 | 9.79 | 1.18 | 0.14 | 0.24 | 0.03 | 91.16 | 14.35 | 1.07 | 91.21 | 9.69 | 1.17 | 0.98 |
| Q26 | 59.89 | 21.88 | 1.64 | 0.20 | 0.01 | 71.76 | 19.92 | 2.42 | 0.23 | 0.06 | 0.00 | 59.89 | 21.88 | 1.64 | 61.60 | 19.91 | 2.41 | 0.57 |
| Q27 | 55.56 | 23.78 | 1.77 | 0.18 | 0.01 | 66.76 | 22.56 | 2.74 | 0.07 | 0.59 | 0.00 | 55.56 | 23.78 | 1.77 | 57.23 | 22.96 | 2.78 | 0.62 |
| Q28 | 82.32 | 19.47 | 1.45 | 0.15 | 0.05 | 88.82 | 14.82 | 1.80 | 0.24 | 0.05 | 0.01 | 82.32 | 19.47 | 1.45 | 83.42 | 14.68 | 1.78 | 0.67 |
| Q29 | 66.78 | 22.39 | 1.67 | 0.28 | 0.00 | 77.94 | 18.00 | 2.18 | 0.24 | 0.05 | 0.00 | 66.78 | 22.39 | 1.67 | 69.32 | 18.62 | 2.26 | 0.41 |
| Q30 | 70.50 | 20.61 | 1.53 | 0.22 | 0.00 | 79.41 | 16.20 | 1.96 | -0.07 | 0.59 | 0.00 | 70.50 | 20.61 | 1.53 | 72.39 | 16.80 | 2.04 | 0.50 |
| Q31 | 59.78 | 23.25 | 1.73 | 0.34 | 0.00 | 68.24 | 20.22 | 2.45 | 0.12 | 0.32 | 0.01 | 59.78 | 23.25 | 1.73 | 62.79 | 21.65 | 2.63 | 0.35 |
| Q32 | 86.03 | 20.19 | 1.51 | 0.05 | 0.52 | 92.75 | 14.91 | 1.80 | 0.15 | 0.22 | 0.01 | 86.03 | 20.19 | 1.51 | 86.42 | 14.78 | 1.78 | 0.88 |
| Q33 | 85.81 | 23.30 | 1.75 | 0.19 | 0.01 | 90.58 | 16.66 | 2.01 | 0.20 | 0.10 | 0.12 | 85.81 | 23.30 | 1.75 | 87.55 | 16.96 | 2.04 | 0.57 |
| Q34 | 76.68 | 24.37 | 1.82 | 0.05 | 0.50 | 76.81 | 21.57 | 2.60 | 0.10 | 0.41 | 0.97 | 76.68 | 24.37 | 1.82 | 77.16 | 21.50 | 2.59 | 0.89 |
| Q35 | 57.63 | 30.59 | 2.30 | -0.12 | 0.11 | 51.45 | 32.34 | 3.89 | -0.32 | 0.01 | 0.16 | 57.63 | 30.59 | 2.30 | 56.17 | 30.85 | 3.71 | 0.74 |
| Q36 | 73.89 | 24.34 | 1.81 | 0.04 | 0.60 | 74.28 | 23.47 | 2.83 | 0.01 | 0.96 | 0.91 | 73.89 | 24.34 | 1.81 | 74.26 | 23.49 | 2.83 | 0.91 |
| SBP | 115.66 | 16.94 | 1.26 | 0.43 | 0.00 | 129.59 | 15.22 | 1.83 | 0.26 | 0.03 | 0.00 | 115.66 | 16.94 | 1.26 | 118.47 | 16.54 | 1.99 | 0.24 |
| DBP | 77.20 | 10.64 | 0.79 | 0.11 | 0.14 | 80.03 | 9.24 | 1.11 | -0.07 | 0.59 | 0.05 | 77.20 | 10.64 | 0.79 | 77.65 | 9.30 | 1.12 | 0.76 |
| HR | 68.93 | 10.80 | 0.80 | -0.15 | 0.04 | 61.19 | 10.10 | 1.22 | -0.19 | 0.12 | 0.00 | 68.93 | 10.80 | 0.80 | 68.29 | 10.07 | 1.21 | 0.67 |
| BMI | 25.83 | 5.81 | 0.43 | 0.04 | 0.61 | 26.53 | 3.69 | 0.44 | 0.00 | 0.97 | 0.35 | 25.83 | 5.81 | 0.43 | 25.92 | 3.69 | 0.44 | 0.91 |
| Gait | 3.10 | 0.47 | 0.03 | 0.19 | 0.01 | 3.26 | 0.61 | 0.07 | 0.24 | 0.05 | 0.03 | 3.10 | 0.47 | 0.03 | 3.13 | 0.60 | 0.07 | 0.64 |
| UAG | 8.58 | 1.91 | 0.14 | 0.20 | 0.01 | 9.35 | 1.77 | 0.21 | 0.08 | 0.51 | 0.00 | 8.58 | 1.91 | 0.14 | 8.73 | 1.81 | 0.22 | 0.59 |

**Supplemental Figure 2.**
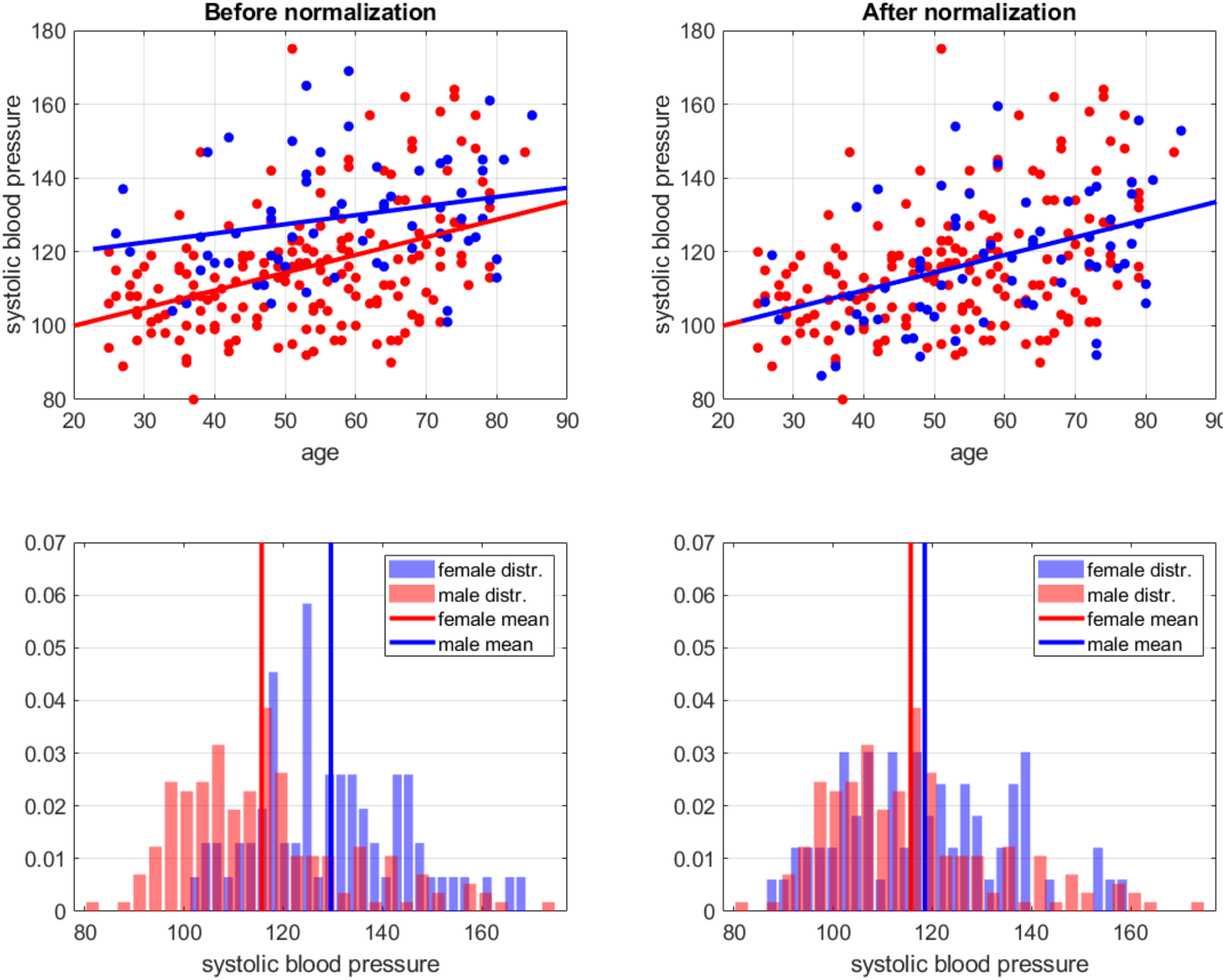
An illustration of the regression-based gender normalization approach. Here, the systolic blood pressure (SBP) increases with age differently for females and males. Thus, aligning the all-age averages would work for middle-aged people but still differ for young and elderly individuals. Instead, we align the regression lines, making the normalization uniform across ages. As shown by the distributions in the bottom panels, this procedure also brings closer the overall averages. Supplemental Tables 1-2 provide the statistics before and after gender normalization for all other measurements used in our analysis.

**Supplemental Figure 3.**
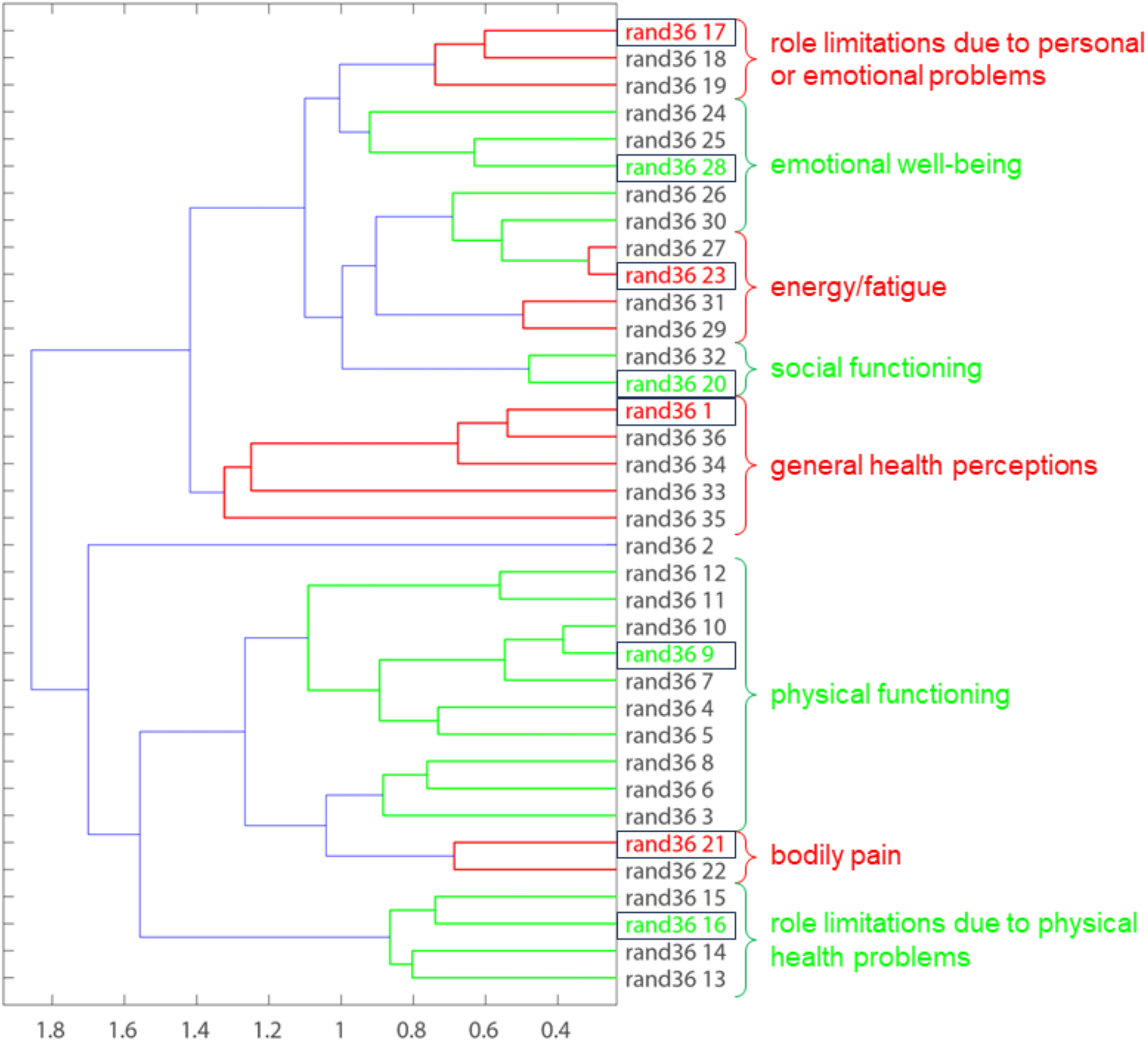
An example of a dendrogram resulted from unsupervised hierarchical clustering of RAND36 survey questions. The x-axis indicates the Euclidean distance between the rows of 36×36 matrix of pairwise correlations. Red and green lines indicate the established (ground truce) grouping of rand36 questions into eight health concepts specified on the right side. The order of questions in this dendrogram coincides with the expected grouping, but a single threshold in the dendrogram distance would not reproduce that grouping. However, as shown in Figure 2, for 33 out of 36 questions, our statistical approach based on 5000 randomized 90% subsets of study participants does produce the correct association with the concept representatives (landmarks) marked here by black boxes.

**Supplemental Figure 4.**
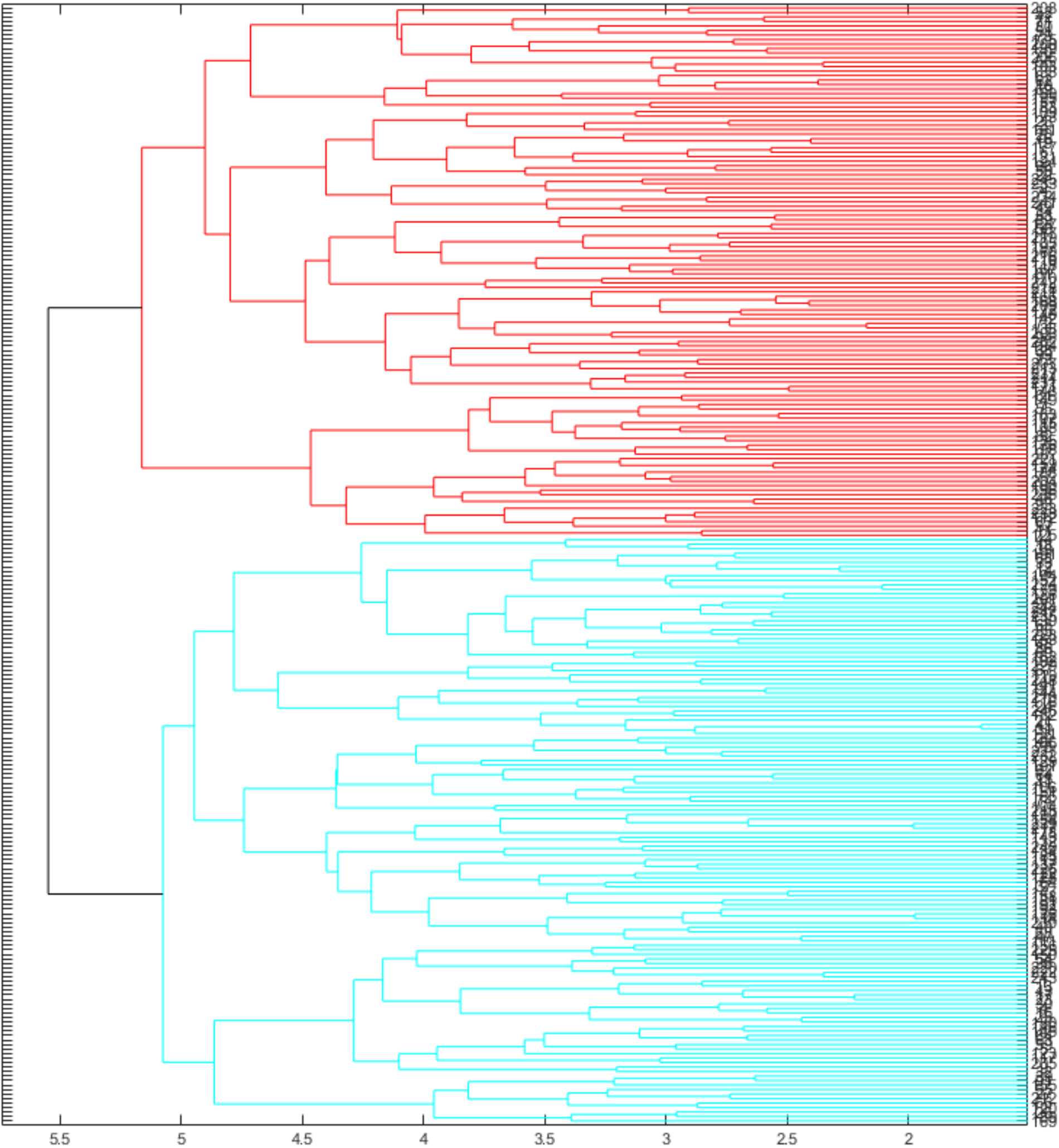
An example of a dendrogram resulted from unsupervised hierarchical clustering of 250 study participants based on clinical lab measurements only. The x-axis indicates the Euclidean distance between the rows in a 250×250 matrix of pairwise correlations. The statistical differences between all measurements in the red and cyan groups are shown in Supplemental Figure 5.

**Supplemental Figure 5.**
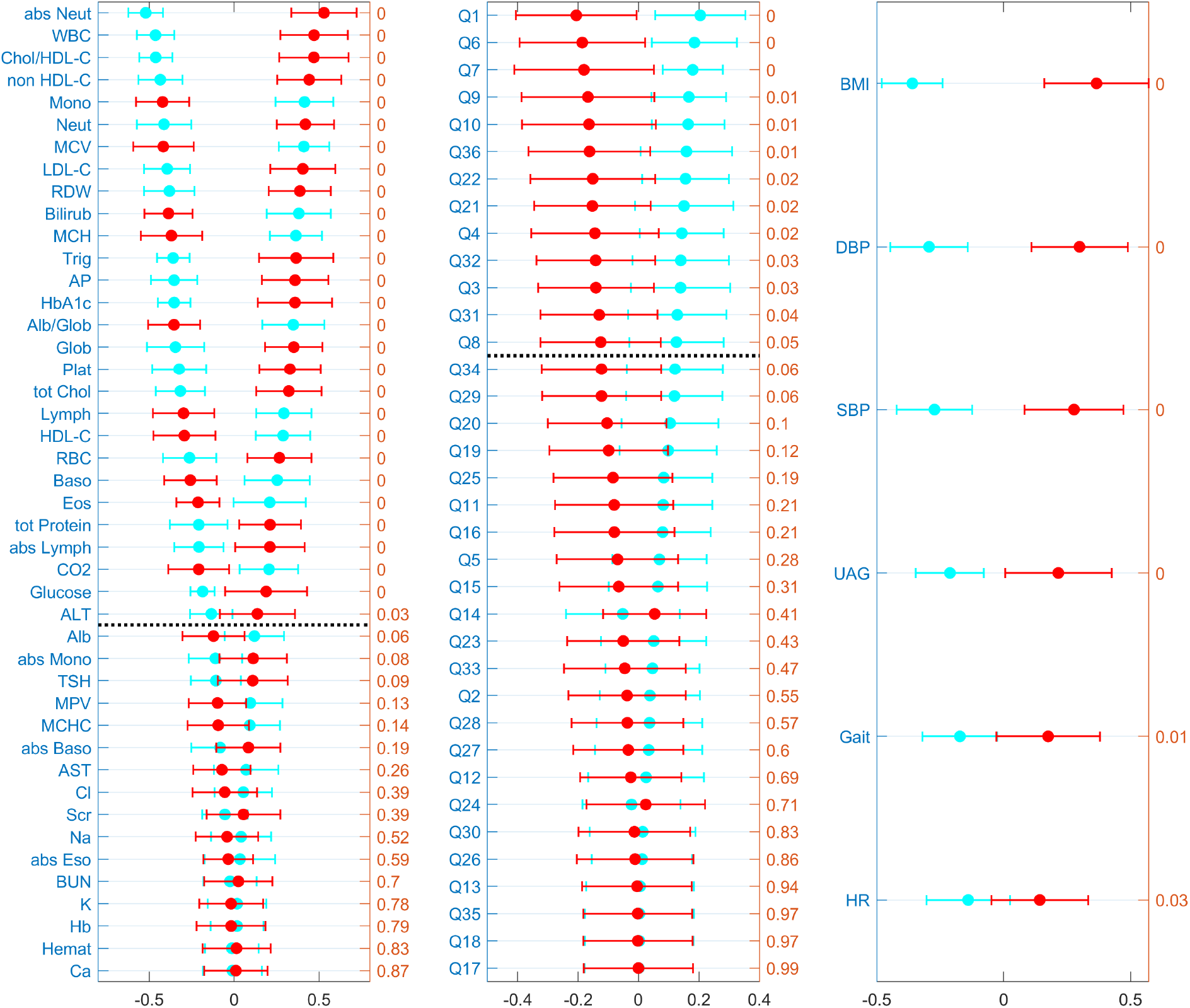
The comparison of mean ± 2 standard error of all measurements in three assessments for two groups of people (red and cyan). The groups are generated by unsupervised hierarchical clustering based on clinical lab measurements as in Supplemental Figure 4. We repeat this calculation 1000 times, every time excluding two randomly selected individuals from the dataset to perturb the clustering and capture the variability of the clustering outcome from the noisy data.

**Supplemental Figure 6.**
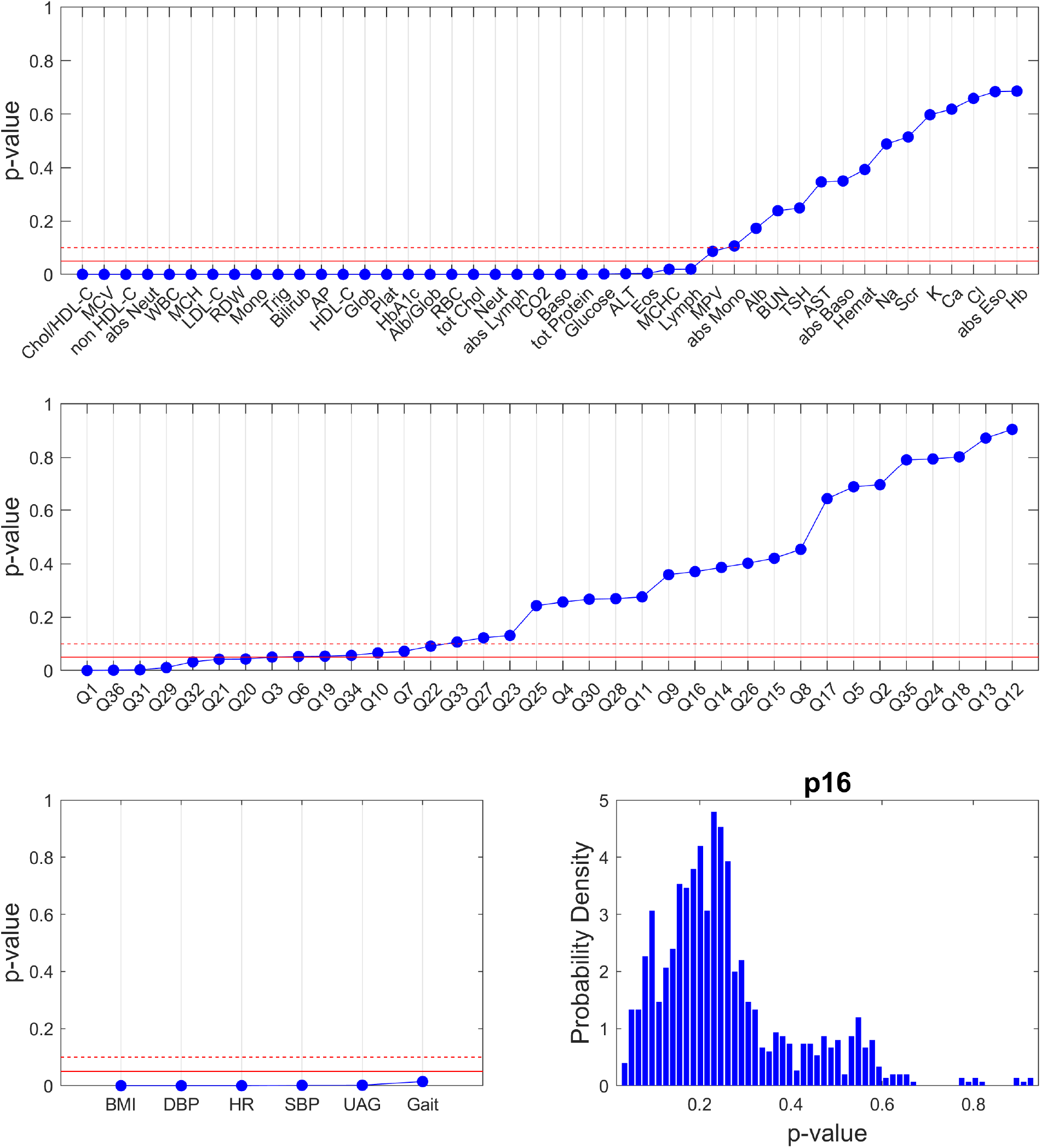
Statistical significance of differences between participant groups based on <u>gender normalized clinical lab</u> measurements using unsupervised hierarchical clustering. The clustering was performed 1000 times; each time excluding two randomly selected people and repeating the procedure for the remaining people. Here p-values validate the null hypothesis that the compared measurements in the two groups have equal mean and variance (two-sample t-test). Blue dots are the median p-values over 1000 randomized repeats for each measure in the dataset. The solid red line corresponds to p-value = 0.05 and the red dashed line to p-value = 0.1. The bottom right graph shows that distribution of p-values for p16 expressions in the resulting clinical lab-based groups.

**Supplemental Figure 7.**
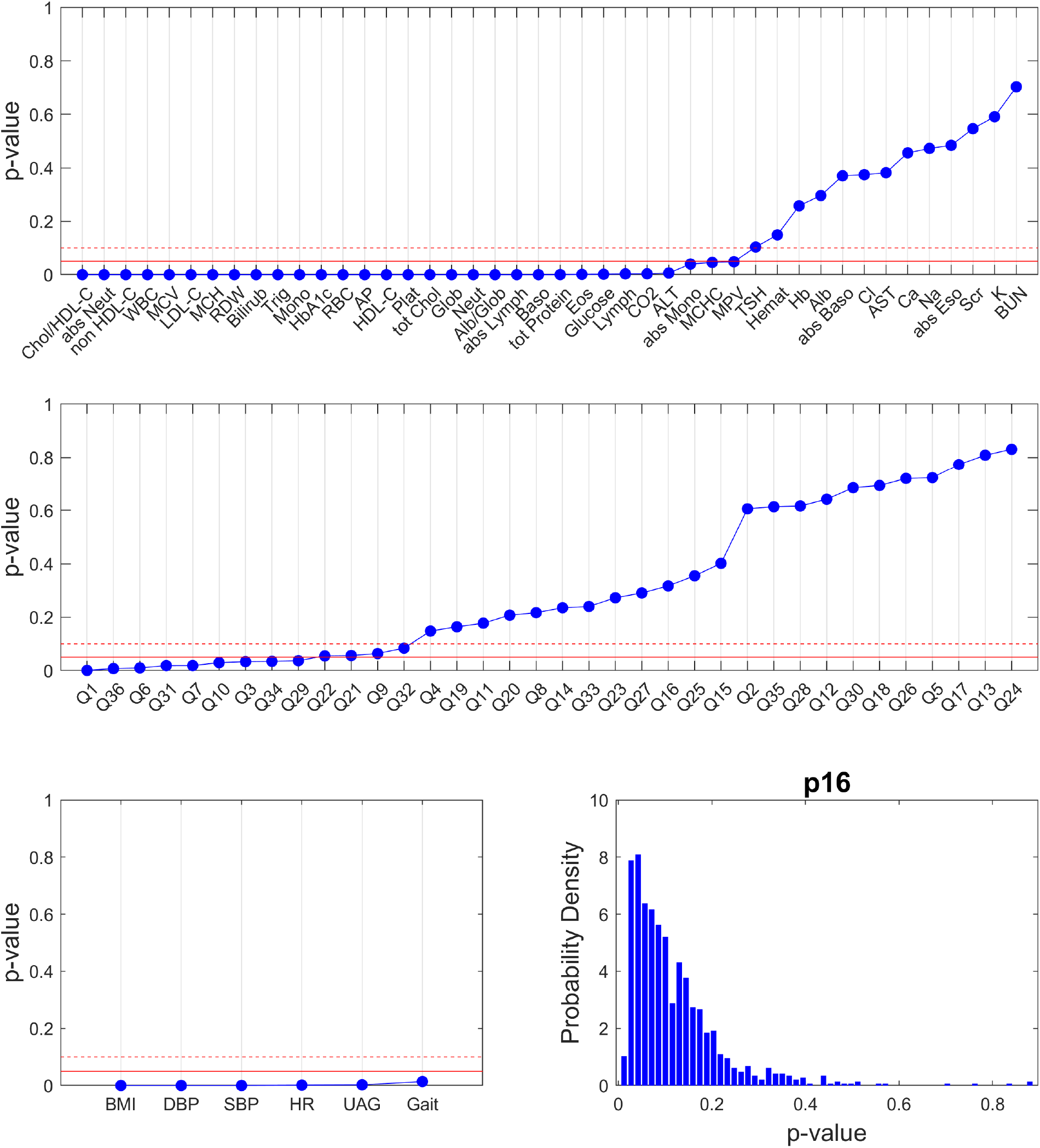
Statistical significance of differences between people grouped based on <u>gender normalized and age corrected clinical lab</u> measurements using unsupervised hierarchical clustering. The clustering was performed 1000 times; each time excluding two randomly selected people and repeating the procedure for the remaining people. Here p-values validate the null hypothesis that the compared measurements in the two groups have equal mean and variance (two-sample t-test). Blue dots are the median p-values over 1000 randomized repeats for each measure in the dataset. The solid red line corresponds to p-value = 0.05 and the red dashed line to p-value = 0.1. The bottom right graph shows that distribution of p-values for p16 expressions in the resulting clinical lab-based groups.

**Supplemental Figure 8.**
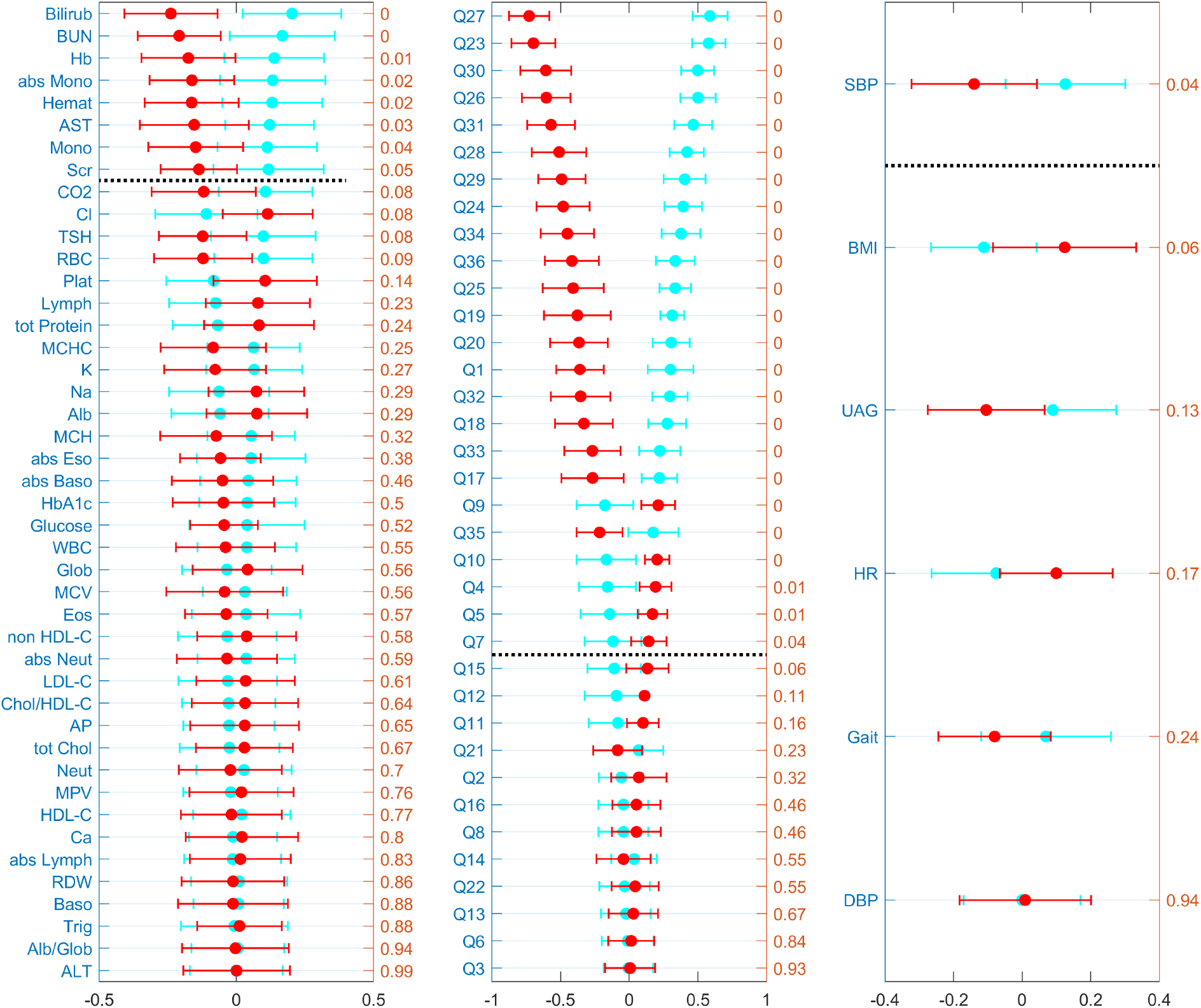
The comparison of mean ± 2 standard error of all measurements in three assessments for two groups of people (red and cyan). The groups are generated by unsupervised hierarchical clustering based on RAND36 survey responses. We repeat this calculation 1000 times, every time excluding two randomly selected individuals from the dataset to perturb the clustering and capture the variability of the clustering outcome from the noisy data.

**Supplemental Figure 9.**
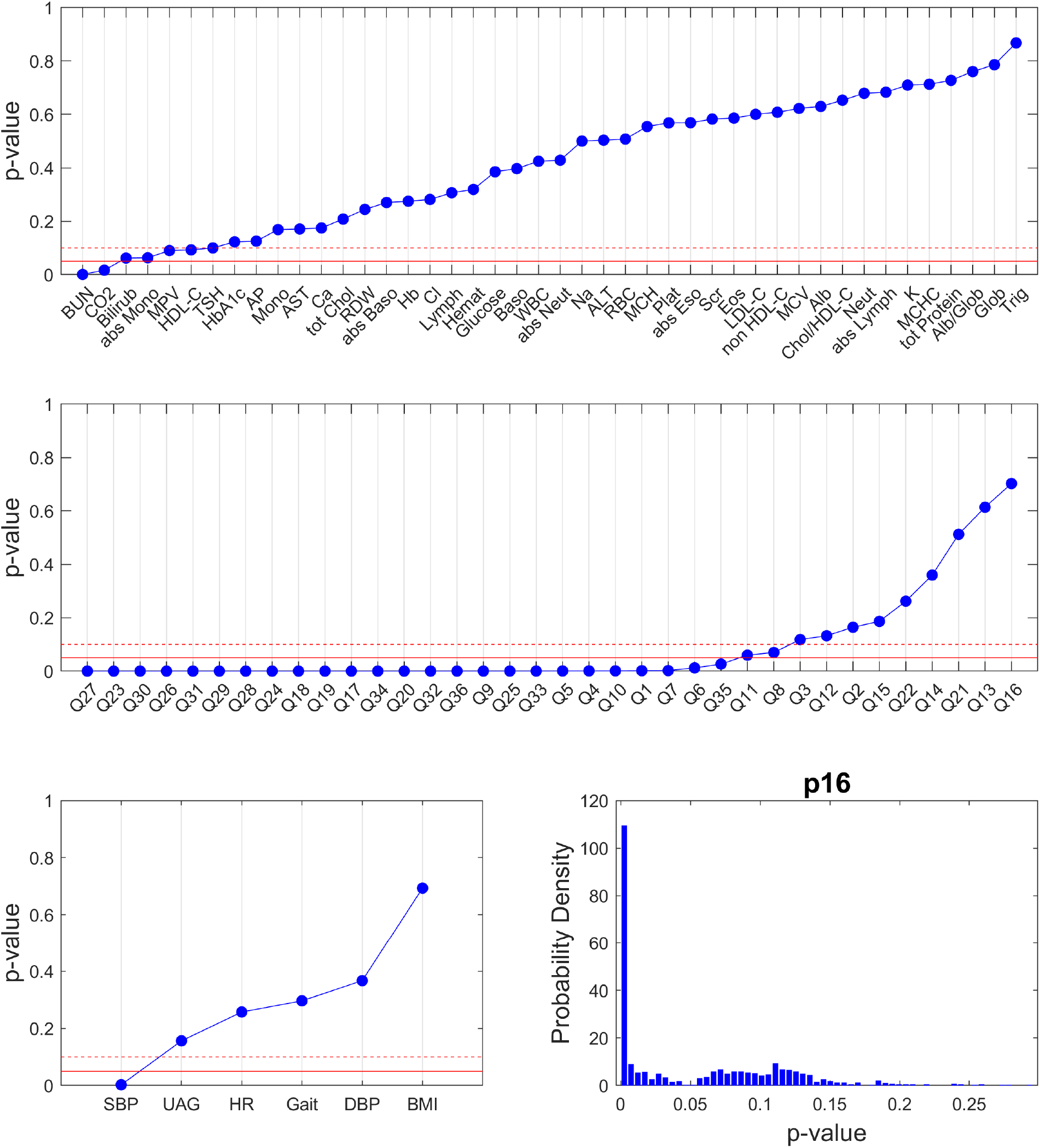
Statistical significance of differences between people grouped based on <u>gender normalized RAND36</u> measurements using unsupervised hierarchical clustering. The clustering was performed 1000 times; each time excluding two randomly selected people and repeating the procedure for the remaining people. Here p-values validate the null hypothesis that the compared measurements in the two groups have equal mean and variance (two-sample t-test). Blue dots are the median p-values over 1000 randomized repeats for each measure in the dataset. The solid red line corresponds to p-value = 0.05 and the red dashed line to p-value = 0.1. The bottom right graph shows that distribution of p-values for p16 expressions in the resulting RAND36-based groups.

**Supplemental Figure 10.**
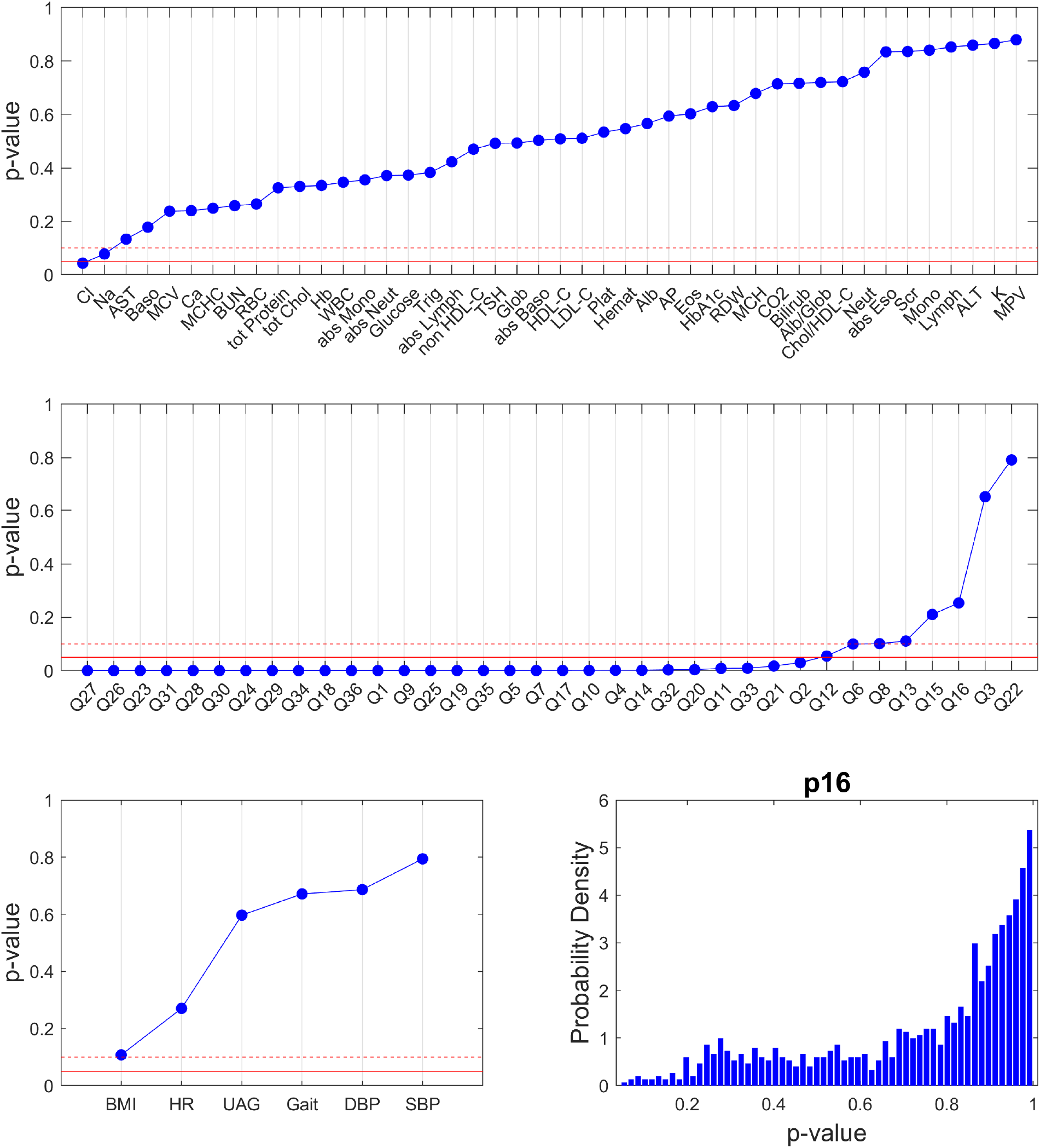
Statistical significance of differences between people grouped based on <u>gender normalized and age corrected RAND36</u> measurements using unsupervised hierarchical clustering. The clustering was performed 1000 times; each time excluding two randomly selected people and repeating the procedure for the remaining people. Here p-values validate the null hypothesis that the compared measurements in the two groups have equal mean and variance (two-sample t-test). Blue dots are the median p-values over 1000 randomized repeats for each measure in the dataset. The solid red line corresponds to p-value = 0.05 and the red dashed line to p-value = 0.1. The bottom right graph shows that distribution of p-values for p16 expressions in the resulting RAND36-based groups.

**Supplemental Figure 11.**
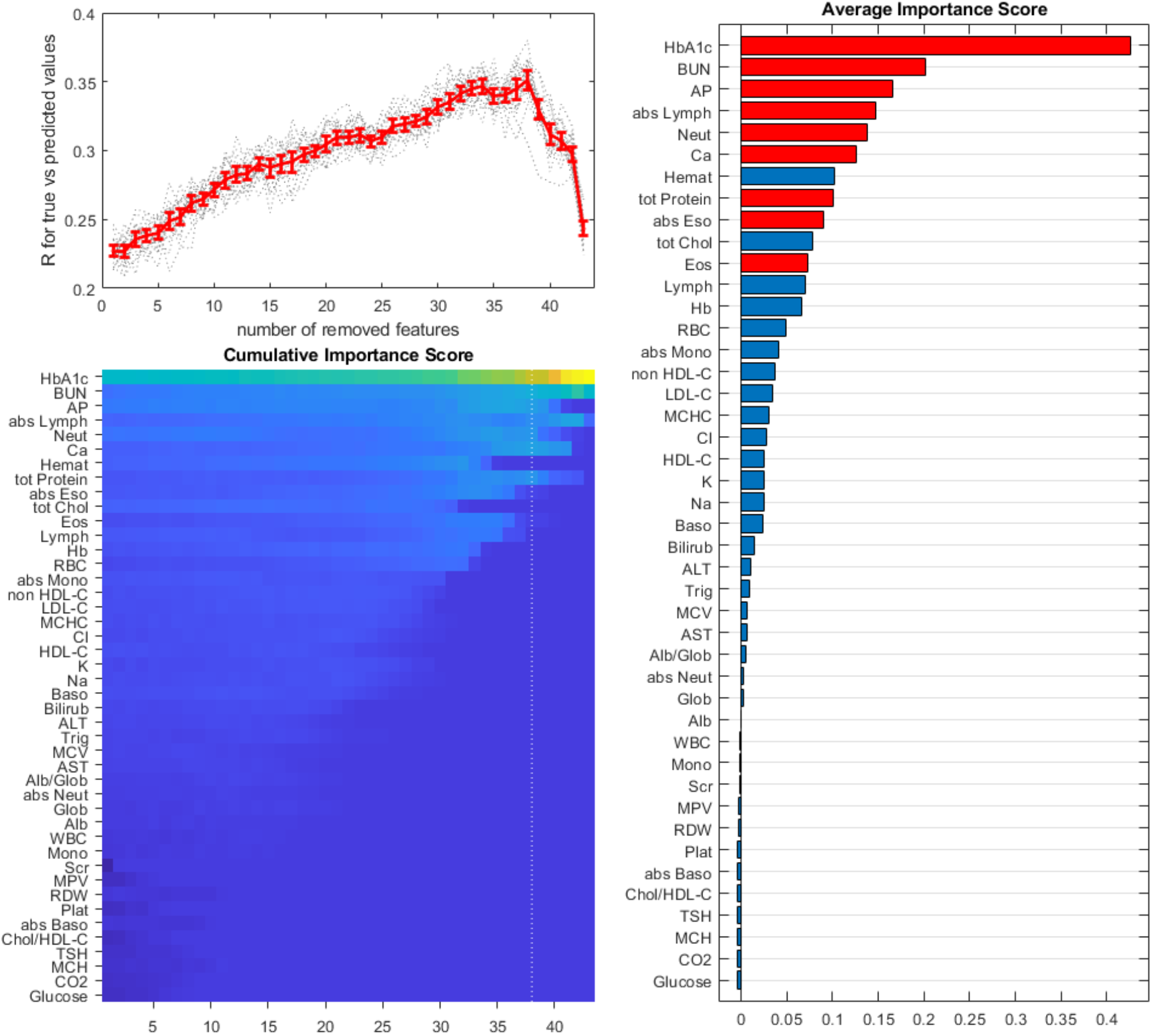
<u>Clinical lab</u> measurements that give the highest prediction accuracy for p16 expression in the <u>gender-normalized</u> dataset. The top left panel shows the correlation coefficient between predicted and true values in the validation set as a function of the number of iteratively removed features with the lowest importance scores. The results for 20 repeats are in gray, and the mean ± 2 standard errors are in red. The bottom left panel shows a colormap of average importance scores over 20 repeats of the feature exclusion protocol. The vertical dotted line corresponds to the exclusion step with the highest prediction accuracy. The right panel shows average importance scores. The red color indicates features presented in the optimal set.

**Supplemental Figure 12.**
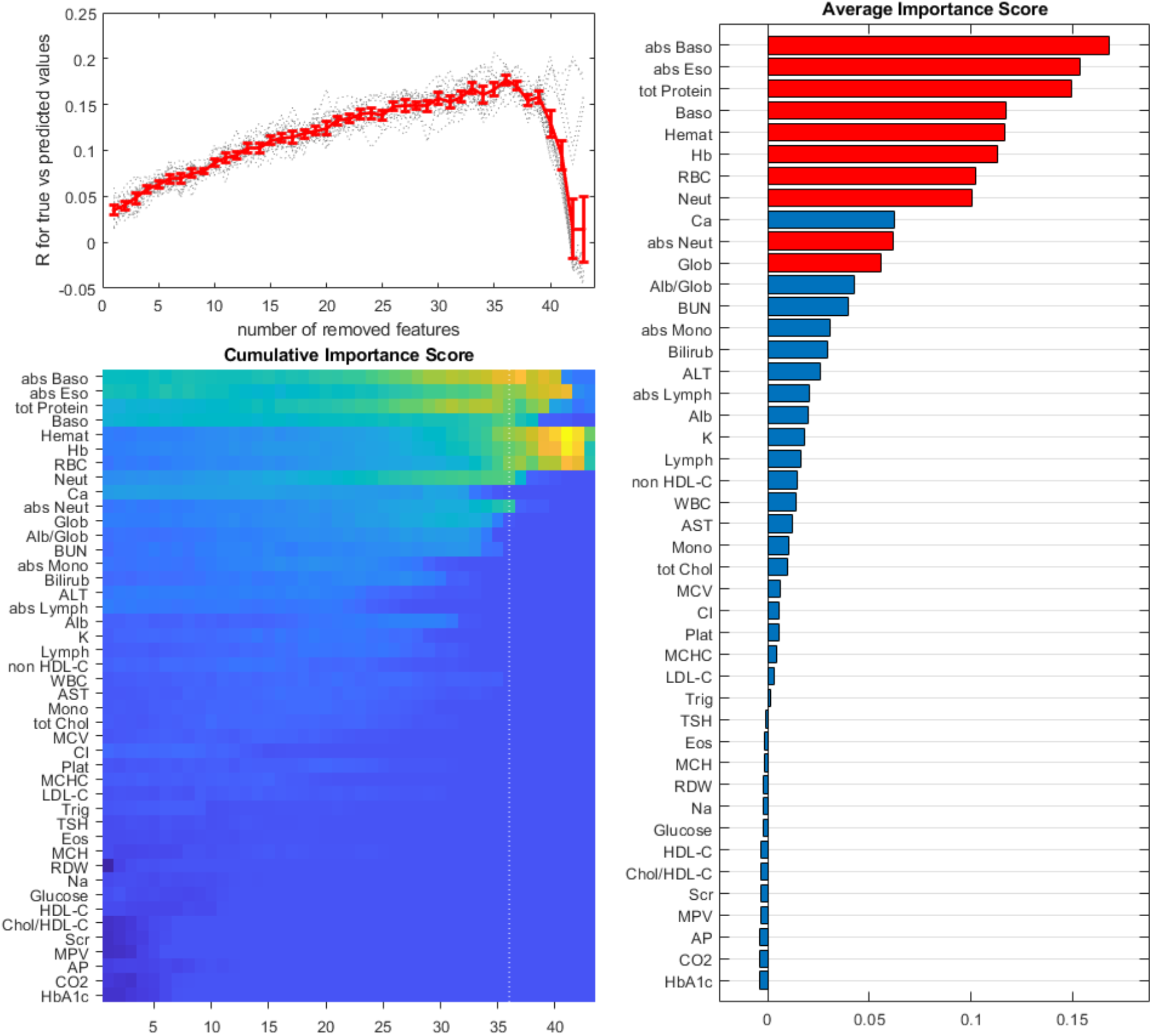
<u>Clinical lab</u> measurements that give the highest prediction accuracy for p16 expression in the <u>age-corrected</u> dataset. The top left panel shows the correlation coefficient between predicted and true values in the validation set as a function of the number of iteratively removed features with the lowest importance scores. The results for 20 repeats are in gray, and the mean ± 2 standard errors are in red. The bottom left panel shows a colormap of average importance scores over 20 repeats of the feature exclusion protocol. The vertical dotted line corresponds to the exclusion step with the highest prediction accuracy. The right panel shows average importance scores. The red color indicates features presented in the optimal set. The optimal prediction accuracy here is much lower than it is without age correction.

**Supplemental Figure 13.**
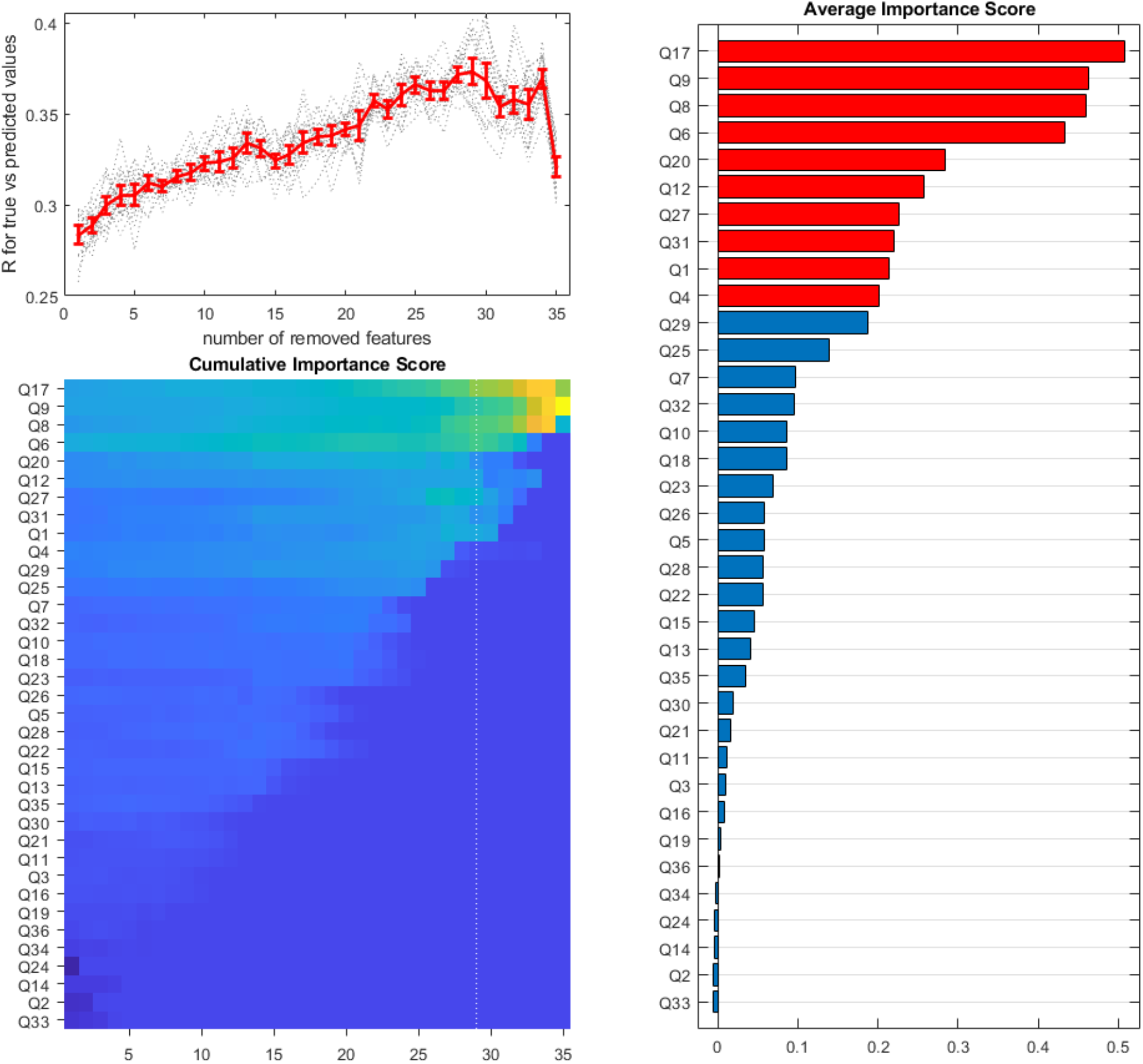
<u>RAND36</u> survey responses that give the highest prediction accuracy for p16 expression in the <u>gender-normalized</u> dataset. The top left panel shows the correlation coefficient between predicted and true values in the validation set as a function of the number of iteratively removed features with the lowest importance scores. The results for 20 repeats are in gray, and the mean ± 2 standard errors are in red. The bottom left panel shows a colormap of average importance scores over 20 repeats of the feature exclusion protocol. The vertical dotted line corresponds to the exclusion step with the highest prediction accuracy. The right panel shows average importance scores. The red color indicates features presented in the optimal set.

**Supplemental Figure 14.**
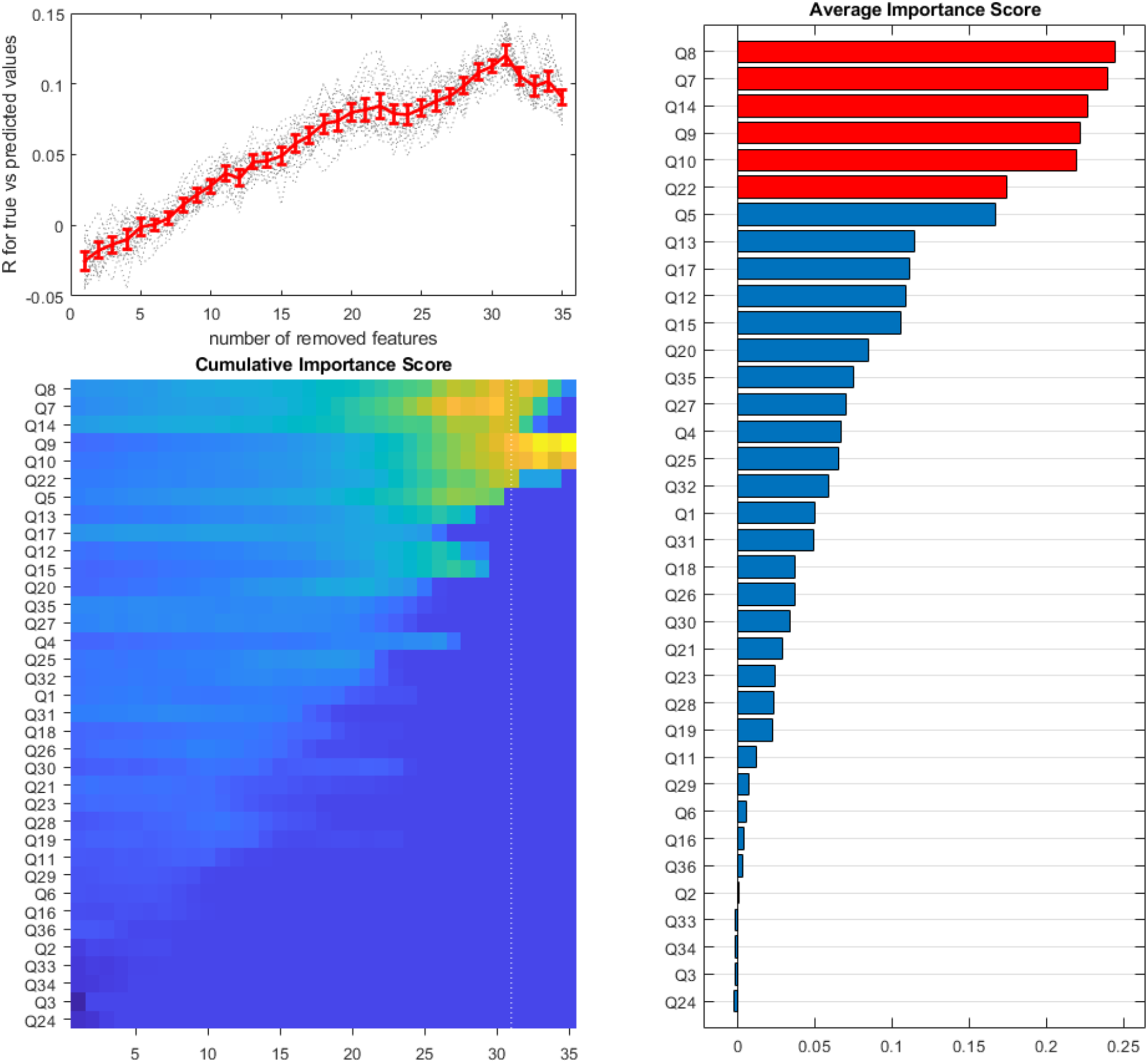
<u>RAND36</u> survey responses that give the highest prediction accuracy for p16 expression in the <u>age-corrected</u> dataset. The top left panel shows the correlation coefficient between predicted and true values in the validation set as a function of the number of iteratively removed features with the lowest importance scores. The results for twenty repeats are in gray, and the mean ± 2 standard errors are in red. The bottom left panel shows a colormap of average importance scores over twenty repeats of the feature exclusion protocol. The vertical dotted line corresponds to the exclusion step with the highest prediction accuracy. The right panel shows average importance scores. The red color indicates features presented in the optimal set. The optimal prediction accuracy here is much lower than it is without age correction.

**Supplemental Table 3:** Optimal feature sets for p16 prediction by clinical labs and RAND36 survey responses. Here **GN** and **AC** stand for ‘gender normalized’ and **‘**age-corrected’ data, respectively.

| Biomarker | Dataset for prediction | Prediction accuracy | Features in the optimal set<br>(in the order of the importance score) |
| --- | --- | --- | --- |
| p16 | Blood | 0.33 | HbA1c, Hemat, Neut, Hb, Lymph, RBC, abs Eso, BUN, AP, Eos, tot Chol, abs Lymph, tot Protein |
| p16 (GN) | blood (GN) | 0.35 | HbA1c, BUN, AP, abs Lymph, Neut, Ca, tot Protein, abs Eso, Eso |
| p16 (AC) | blood (AC) | 0.18 | abs Baso, abs Eso, tot Protein, Baso, Hemat, Hb, RBC, Neut, abs Neut, Glob |
| p16 | RAND36 | 0.29 | Q8, Q31, Q29, Q1, Q17, Q9, Q3, Q27, Q20, Q25, Q26, Q6 |
| p16 (GN) | RAND36 (GN) | 0.37 | Q17, Q9, Q8, Q6, Q20, Q12, Q27, Q31, Q1, Q4 |
| p16 (AC) | RAND36 (AC) | 0.12 | Q8, Q7, Q14, Q9, Q10, Q22 |

**Supplemental Table 4:** Formulas for clinical lab-based and RAND36-based indexes that have the highest correlation coefficients with p16 expression.

| Composite index | Correlation coefficient | Percent improve | Formula |
| --- | --- | --- | --- |
| $S_{\text{labs},p16}$ | 0.39 | 215 | tot Chol + HDL-C + LDL-C + non HDL-C + BUN + Scr + Na + K + Ca + Glob – Alb/Glob + AP + HbA1c + Hb + Hemat + MCV – MCHC – MPV – abs Lymph – abs Mono – abs Eso – abs Baso + Neut – Lymph |
| $S_{\text{rand36},p16}$ | 0.33 | 134 | – Q3 – Q8 + Q17 + Q29 |

